# Who should be first in line for the COVID-19 vaccine? Surveys in 13 countries of the public’s preferences for prioritisation

**DOI:** 10.1101/2021.01.31.21250866

**Authors:** Raymond Duch, Laurence S J Roope, Mara Violato, Matias Fuentes Becerra, Thomas Robinson, Jean-Francois Bonnefon, Jorge Friedman, Peter Loewen, Pavan Mamidi, Alessia Melegaro, Mariana Blanco, Juan Vargas, Julia Seither, Paolo Candio, Ana Gibertoni Cruz, Xinyang Hua, Adrian Barnett, Philip M Clarke

**Affiliations:** University of Oxford; Universidad de Chile; University of Durham; Toulouse School of Economics; Universidad de Santiago de Chile; University of Toronto; Center for Social and Behavioral Change; University of Bocconi; Universidad del Rosario; University of Melbourne; Queensland University of Technology

## Abstract

How does the public want a COVID-19 vaccine to be allocated? We conducted a conjoint experiment asking 15,536 adults in 13 countries to evaluate 248,576 profiles of potential vaccine recipients that varied randomly on five attributes. Our sample includes diverse countries from all continents. The results suggest that in addition to giving priority to health workers and to those at high risk, the public favours giving priority to a broad range of key workers and to those on lower incomes. These preferences are similar across respondents of different education levels, incomes, and political ideologies, as well as across most surveyed countries. The public favoured COVID-19 vaccines being allocated solely via government programs, but were highly polarized in some developed countries on whether taking a vaccine should be mandatory. There is a consensus among the public on many aspects of COVID-19 vaccination which needs to be taken into account when developing and communicating roll-out strategies.

## Introduction

How to allocate a COVID-19 vaccine is one of the most important decisions currently facing every government. There has been an unprecedented race to develop a vaccine. At the time of writing, 58 vaccine candidates are currently undergoing human trials (*1*). Several vaccines have been shown to be safe and highly effective (*2*) and countries are starting to approve their use (*3*).

It has been recognised that, in many countries, public confidence in vaccination is fragile, and that policies for prioritizing vaccine allocation need to be seen as both equitable and evidence-based (*4*). Ethical frameworks have been suggested for the allocation of scarce vaccine supplies between countries (*5*). The WHO has developed a values framework based on twelve objectives and six principles (Human Wellbeing, Equal Respect, Global Equity, National Equity, Reciprocity, Legitimacy). Importantly, the WHO does not provide any guidance on the order of importance of either the principles or the objectives. (*6*) Constraints on timely supply of vaccines mean that it is likely that it will not be possible to secure all of the objectives simultaneously. The WHO Strategic Advisory Group of Experts on Immunization (SAGE) has recently proposed a roadmap which prioritizes health workers and older adults. (*7*)

At a national level, governments are having to rapidly develop guidelines to prioritize access to a vaccine. Based on a survey of governments’ vaccine allocation policy plans, conducted in early December 2020, Table 1 indicates that there is considerable diversity across countries in the groups being prioritized. While prioritization of health workers and the clinically vulnerable appears universal, there is little consensus on which other groups to prioritize. The UK prioritization strategy is largely aged-based, starting with the oldest age categories followed by the clinically vulnerable (*8*). No other criteria will be employed until after everyone over 50 and/or with underlying health conditions has been vaccinated. In contrast, an expert committee in France has recommended prioritizing workers who have contact with the general public including shop workers, school staff, transport staff and hospitality workers. In the US, the Centres for Disease Control and Prevention will decide whether to prioritize essential workers including school staff, police, grocery workers, and bus drivers or adults over 65 and those of any age who have high-risk medical conditions. (*9*) Chile appears to be planning yet a different strategy, prioritising health care workers, other essential workers and teachers. In sum, there is substantial variation in who can get a vaccine and when.

**Table 1:** Criteria proposed or used to prioritize COVID vaccine allocation by country

|  | Age | COVID-19 |  |  | Occupation |  |
| --- | --- | --- | --- | --- | --- | --- |
|  |  | Transmission | Vulnerability | Essential<br>infrastruc-<br>ture | Health/social<br>care | Education/child<br>care |
| Australia | x | x | x | x | x |  |
| Brazil | x |  | x | x | x | x |
| Canada | x | x | x | x | x |  |
| Chile |  |  |  | x | x | x |
| China | x | x | x | x | x |  |
| Colombia | x |  | x |  | x |  |
| France | x | x | x | x | x |  |
| India | x |  |  | x | x |  |
| Italy | x |  | x |  | x |  |
| Spain |  |  | x |  | x |  |
| Uganda |  |  | x |  | x |  |
| UK | x |  | x |  | x |  |
| USA | x |  | x | x | x |  |

Many health technology assessment (HTA) agencies involve the public in decisions (*10*). Such processes have, however, been largely absent from the development of guidelines for COVID-19 vaccine prioritization. While HTA agencies often involve patient representatives (*11*), wider input including the use of citizen juries (*12*) and surveys of public preferences (including use of conjoint methods (*13*)) has long been advocated. While there have been calls for the public to have a say in COVID-19 vaccine priority setting (*14*), to date empirical evidence on public preferences has been very limited (*12, 15, 16*).

Beyond priority setting, governments will need to consider a number of vaccine policy measures that may, or may not, be seen by the general public as equitable and fair. Some governments are debating whether citizens will be able to purchase a COVID-19 vaccine from private providers. For example, the Australian government has indicated the potential for private sales of vaccines, while in India, one vaccine is expected to go on sale within a few months. (*17*) On the other hand, in many countries there are no plans for the private sale of COVID-19 vaccines. (*18*)

Governments are also considering whether they should make COVID-19 vaccination mandatory. There are strong ethical arguments (*19*) for forms of coercion in public health to deal with the externalities that arise from infectious diseases (i.e. those that refuse vaccination not only put themselves at risk, but increase the risk to others). While a recent international survey on factors that could influence potential COVID-19 vaccine uptake indicated that employer-mandated vaccination would decrease the likelihood of use, governments already have in place policies to provide strong incentives for uptake of existing vaccines. The merits of some form of mandating have already been subject to considerable public discussion. (*20*). Nonetheless, we do not know whether, where and to what extent, mandates are supported by the general public.

The successful roll out of COVID-19 vaccines will depend on high uptake. An important element of this successful roll out is a public that views the adopted prioritization system as fair and equitable. If this is not the case, for whatever reasons, governments risk the types of public resistance and polarization that occurred in some countries regarding the wearing of masks (*21*). It also risks the creation of vaccine black markets that would threaten the safety and fairness of the vaccination campaign. To accomplish these goals governments should seek evidence of the public’s opinions and preferences regarding the groups to be prioritized, public versus private distribution channels and mandatory requirements to be vaccinated. This information can aid in the design of better policies and the implementation of successful communication campaigns, both of which would help ensure a successful COVID-19 vaccination program (*22*).

## 1 Study Design

To provide an evidence based understanding of public opinions on key aspects of vaccine allocation, we undertook a large scale online public opinion survey in 13 countries. Quota sampling was employed in order that the survey matched the demographic margins from the populations of each country.^1^ The survey included a conjoint experiment to identify preferences for different vaccine prioritization schemes. Conjoint survey experiments are frequently employed to identify the importance individuals attribute to different features or characteristics of choices (*23*). Examples include environmental migrants (*24*), asylum seekers (*25*) and migration destinations (*26*). (*27*) employed conjoint experiments that generated 40 million decisions to determine the ethical principles the public thinks should guide self-driving cars.^2^ In the case of policy-oriented survey experiments, evidence suggests that the weights given to attribute characteristics in conjoint survey experiments map closely to the actual policy choices made by the population (*29*).

In our conjoint experiment each of the 15,536 subjects made eight binary choices over hypothetical vaccine recipients (a total of 124,288 pair-wise comparisons) that randomly varied on five attributes: occupation, age, transmission status (risk of contracting and transmitting the virus), risk of death from COVID-19, and income.^3^ Table 1 suggests that these five attributes have played particularly important roles in the vaccine allocation policies being considered by our sample of countries.

## 2 Global COVID-19 Vaccine Allocation Priorities

We estimated the importance of specific characteristics of vaccine allocation priorities with logistic regression (with standard errors clustered by participant). For each pair-wise choice we regressed the participant’s binary decision on dichotomous variables representing the attribute values of the five vaccine allocation attribute variables. Table 6 in the Supplementary Materials presents the logistic regression results. Figure 1 reports the coefficients, along with their confidence intervals, from this logistic regression. We are interested in the relative effects of the attribute-levels in our models, and the logistic coefficients are sufficient to demonstrate the difference in relative magnitude and the direction of any causal effects within and between attributes. Readers should note that these coefficients should not be directly interpreted as the marginal effects on the probability of choosing a vaccine recipient.^4^ The reference categories for the conjoint attributes are the neutral categories included as dots with coefficient zero in Figure 1.

**Figure 1:**
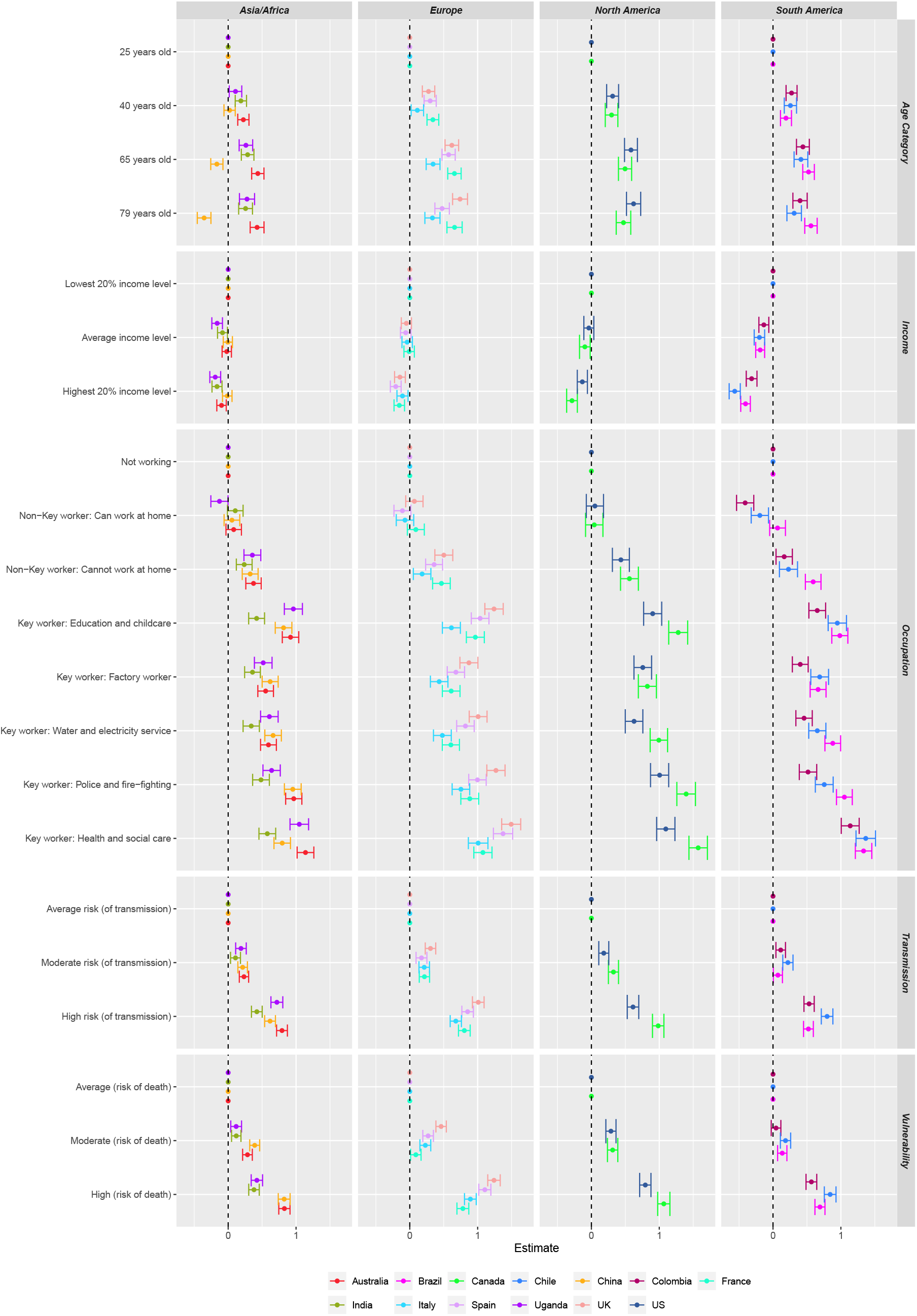
Vaccine Candidate Decisions by Country: Logistic coefficients are reported for attribute values. 95% confidence intervals are shown for each point estimate, clustered by subject.

The individual country conjoint results are organized by three regions in Figure 1. There is no evidence in any of the 13 countries of respondents treating all potential vaccine recipients equally. With respect to each of the five priority attributes there is evidence that the global public favors some profile attributes over others. Moreover, the pattern of coefficient values across our sample of 13 countries is quite similar. The global public exhibits a surprising consensus on which population segments should have priority for a COVID-19 vaccine.

*Age* matters. Respondents in virtually all countries favor vaccine candidates with age profiles greater than the young, 25 year-old, reference category. And there is evidence in a number of countries suggesting that the two oldest age categories (the 65 and 75 year old profiles) were favored over the younger 25 and 40 year old profiles. The one exception is China, where the older 65 and 75 year old profile attributes were less preferred than the younger, 25 and 40 year old, profile attributes. This distinct preference in China for younger vaccine recipients might be a function of the very young age composition of the China sample, which is heavily skewed towards younger participants.

For a majority of the 13 countries in our study, the *income* attributes of the potential vaccine recipients affects allocation preferences. The reference category here is the lowest quintile of the income distribution. In most middle and low income countries (Brazil, Chile, Colombia, India and Uganda), respondents exhibit lower preferences for potential vaccine recipients in the average and high income categories. We see a similar, although somewhat less pronounced, pattern for the U.S., Canada, China, Australia, and European countries. It is worth pointing out that redistributive vaccine allocation preferences seem particularly salient in those countries with higher levels of income inequality.

Vaccine allocation preferences related to *occupation* are virtually identical across all the sampled countries. The reference category here is “not working”. In all of the countries, respondents accorded similar vaccine priority to the “not working” profile attribute and the “non-key worker able to work at home” attribute. The “non-key workers unable to work at home” attribute had a significantly positive impact on profile selection (relative to the “not working” reference category). All of the “key worker” occupational attributes had a positive effect on vaccine recipient selection in all sampled countries. This “key-worker” result is very much consistent with the recent vaccine allocation priority plans announced by the governments of our sampled countries (see Table 1).

As Table 1 indicates, many of the governments of our sampled countries have prioritized COVID-19 vaccination for individuals who are at *high risk of death* from the virus and, to a lesser extent, those at *high risk of contracting and transmitting the virus*. Similarly, in Figure 1, we see that vaccine allocation profiles with “high risk of COVID-19 death” and high risk of “COVID-19 contracting and transmitting” were significantly more likely to be selected by respondents in all 13 country samples.

Segments of the population may display quite different views on vaccine allocation priorities. Figure 2 explores this possible heterogeneity by breaking out the main conjoint analysis by age, income and left-right political self-identification. Overall, the effects of the attributes were broadly similar across the different subgroups. As to which vaccine recipient attributes should be prioritized, there is a general consensus among those identifying with the left and the right, young and old, less and more highly educated, and richer and poorer citizens. In the Supplementary Materials (see Figure 7) we conducted additional sub-group analyses indicating that women and men, high and low educated, and those intending and not intending to be vaccinated all agree on the vaccine recipient attributes that should be prioritized. These are a diverse set of possible sources of heterogeneous treatment effects. This absence of heterogeneity here suggests that preferences regarding vaccine priorities are not affected by self-interest, political partisanship, vaccine hesitancy, or educational attainment. Instead, they reflect broad, societal consensuses on who should be vaccinated.

**Figure 2:**
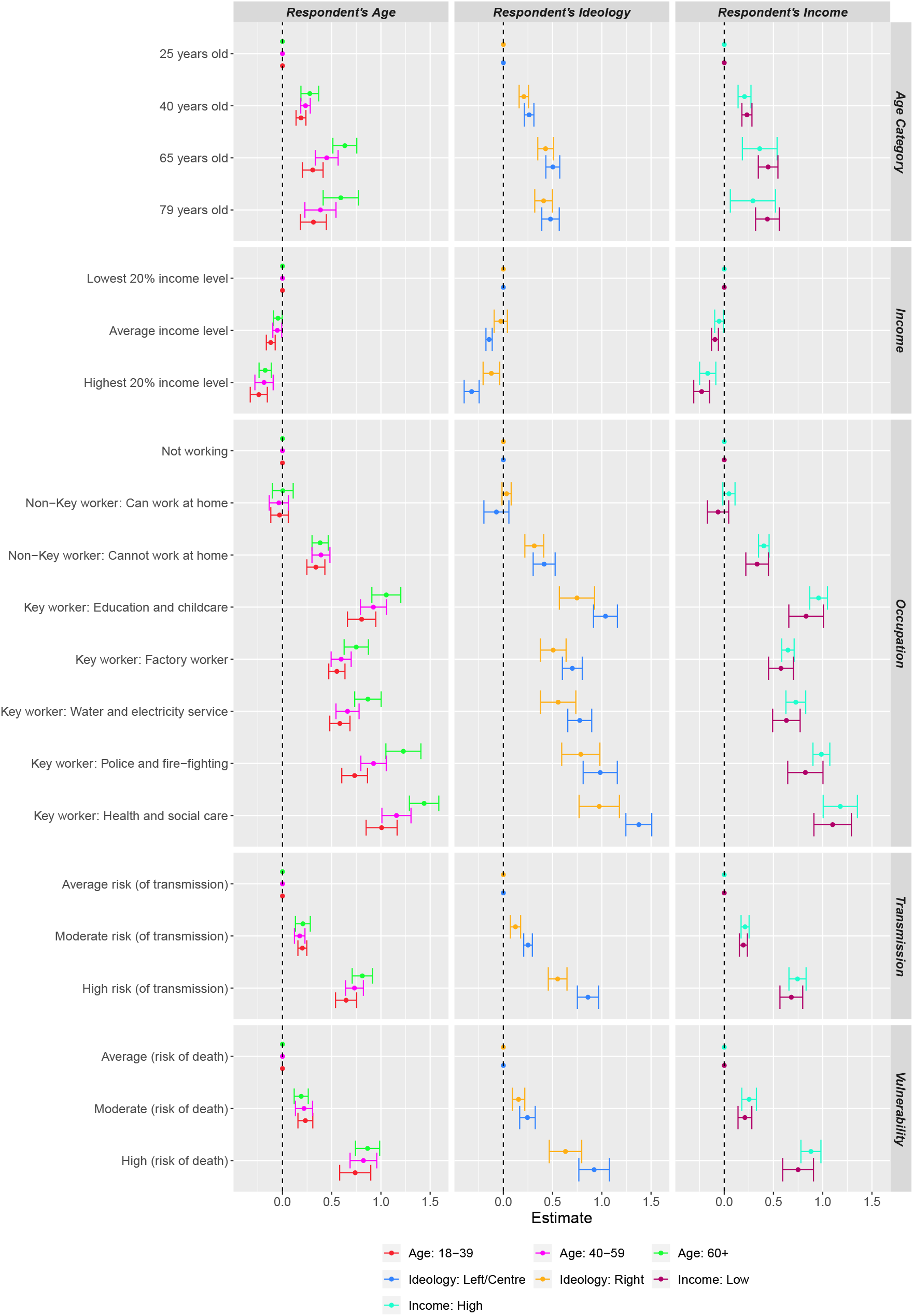
Vaccine Candidate Decisions by Age, Income and Left-Right Self-Identification: Logistic coefficients are reported for attribute values. 95% confidence intervals are shown for each point estimate, clustered by country.

## 3 Global COVID-19 Allocation Mechanisms

Implementing COVID-19 vaccination programs will be challenging. The COVID-19 vaccines are a public good and the general public has expectations regarding how this public good should be provided; specifically, what vaccine allocation mechanisms are appropriate or acceptable. The global survey included questions measuring preferences for how governments should implement the vaccination program. The results, summarized in Figures 3, suggest that the public is not indifferent to the allocation mechanisms put in place.^5^

**Figure 3:**
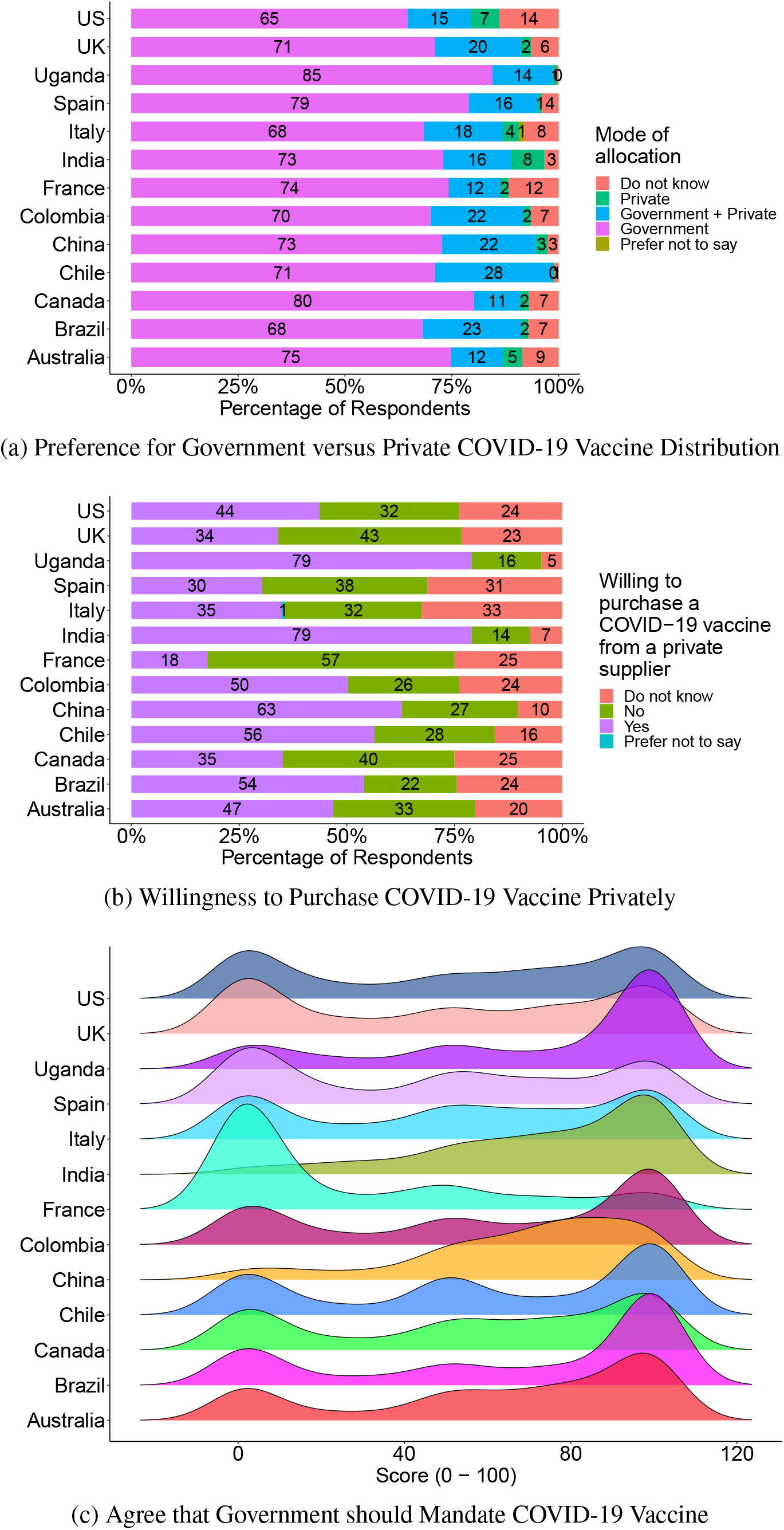
Allocation Mechanism

First, there is evidence of a global consensus on the important role of government distribution. Respondents in the survey were asked whether the COVID-19 vaccine should be made available through government distribution alone; government distribution and private sale; or only private sale. Panel A in Figure 3 confirms that a very large majority of the public believes the government should assume the lead role in the distribution of COVID-19 vaccines. Moreover, this result is consistent across each of the 13 countries; at least two thirds of the population in each country said that distribution should only be available via government schemes. Nevertheless, in most countries there are still substantial percentages of the population who feel there should be at least some role for private distribution; this ranges from 13 percent in Canada to 28 percent in Chile.

While a majority in all countries think COVID-19 vaccine distribution should be solely via government schemes, there is evidence that a large proportion would be willing to pay for a vaccine if it were available privately. We asked respondents “If a COVID-19 vaccine was also available for private purchase and you could receive it immediately [rather than wait six months] would you considering buying it.” Panel B in Figure 3 suggests that roughly half of our global sample would be willing to purchase the COVID-19 vaccine on the private market, ranging from 18 percent in France to 79 percent in India and Uganda. The low and middle income countries are particularly enthusiastic about purchasing the vaccine on the private market (while at the same time strongly favoring government provision).

There was no consensus on whether the COVID-19 vaccine should be mandatory, either globally or within national borders. We asked respondents to indicate on a scale from 0 (very much disagree) to 100 (very much agree) how much they agreed or disagreed with the statement “government should make the COVID-19 vaccine mandatory for everybody.”

As panel C in Figure 3 indicates, opinion overall is skewed toward making COVID-19 vaccination mandatory. But there is evidence of some polarization. In a number of countries there is substantial clustering of responses at both ends of the scale. About 24 percent of our global sample were strongly opposed to mandatory vaccination, while about 38 percent were strongly in favour. There was variation amongst countries. In France, there was a broad consensus of opinion strongly opposing mandatory vaccination (about 60 percent opposed mandatory vaccination). Opinion was highly polarized in the US and UK, with the majority of people either strongly opposed to mandatory vaccination, or strongly supportive. Opinion was also somewhat polarized, with little middle ground, in Australia, Brazil, Chile and Colombia - but with a much larger cluster supporting mandatory COVID-19 vaccination. In China, India, and Uganda, very few people were strongly opposed to mandatory vaccination and the majority were strongly supportive.

## 4 Discussion and Conclusion

This study undertook a representative online survey of 15,536 members of the general public in a diverse range of countries to understand preferences and opinions regarding the allocation of a COVID-19 vaccine. To elicit preferences we undertook a large-scale conjoint experiment in which respondents were asked to evaluate profiles of candidate recipients for a COVID-19 vaccine. The profiles varied on a set of randomly assigned attributes. We found that the public would prioritize people for vaccination based on a broad range of factors. Not surprisingly, these include features directly related to contracting COVID-19 or developing severe symptoms, such as age, vulnerability and risk of transmission. Notably, however, the public would also prioritise according to what might be deemed more economic factors, including low income groups and quite a wide range of non-health related key occupations (e.g. teachers) and non-key workers that cannot work from home. While there is substantial variation in COVID-19 vaccine allocation policies in our sample of countries (see Table 1), public preferences for prioritization appear to be largely consistent across countries and larger regions. Moreover, when we estimate the conjoint model for sub-groups in the samples, we find that preferences are consistent across a range of respondent characteristics such as age and income (Figure 2). Preferences for prioritization also do not vary by political leanings, suggesting that if governments implement policies in line with these public views they should not be politically polarizing.

Conjoint methods have been widely used to understand preferences for other types of health care, including influenza vaccination, and have been shown to mimic real world decision making. However, it is important to note that our conjoint experiment is largely intended to capture preferences for prioritizing the access of others. Our analyses of heterogeneity indicate that in many instances respondents are willing to prioritize individuals that do not share their own characteristics. For example, those aged over 65 were found to have strong preferences for prioritising many types of essential workers. It is difficult to disentangle the degree to which these preferences are due to altruism, or to a desire to reduce the likelihood of being infected through interaction with unvaccinated individuals; or more broadly a desire to choose an optimal vaccination program that would help end the pandemic and bring life back to normal sooner.

A limited number of studies have examined COVID-19 vaccine preferences at a national level. For example, a recent survey of the Belgian population (*16*) found a preference for vaccination of essential workers, those more likely to transmit the virus and those at high risk. Unlike our study, it did not find any preference for allocation to those aged over 60. An advantage of conducting comparative international studies of preferences and opinions is the ability to provide evidence on the degree to which public views are consistent across geographically, economically and culturally diverse countries. While there have been efforts to understand preferences for different methods of social distancing (*31*) and the likely uptake of a COVID-19 vaccine (*32*), we have not found other comparable international surveys that can help inform COVID-19 vaccine allocation.

Our study also has important implications for vaccine prioritization policies. Importantly, in the countries in our sample, there appears to be much greater heterogeneity in national policies (see Table 1) than in public preferences. While current policies include some of the groups that the public think should be given priority (e.g. those at high risk of mortality), there are only a minority of countries that give priority to vaccinating groups of key workers (such as teachers). Further, our study indicates that the public feel that a broader set of economic factors should be taken into account in prioritization policies - these include low income groups and non-essential workers that cannot work from home. With regard to the former, while some countries have stressed the importance of ensuring good vaccine uptake among deprived groups, we are unaware of any countries that are explicitly targeting people according to levels of income or other markers of deprivation. (*33*)

Regarding the mode of allocation, there is a remarkably consistent preference for government-only allocation across all countries (Figure 3). Nevertheless, the public will pay for a COVID-19 vaccine if it does become privately available. We saw in Figure 3 that in many countries a significant proportion indicated a willingness to purchase it on the private market in order to receive the vaccine faster. The highest proportion of those willing to purchase privately is in low and middle income countries such as Brazil, Chile, India and Uganda. Given the emergence of companies that are gearing to supply privately (*17*), understanding implications for both coverage and affordability should be a priority. If COVID-19 vaccines are to be allocated privately, it will be essential to develop policies to ensure that private allocation does not jeopardize a country’s ability to acquire the doses necessary for government-managed vaccination campaigns. Ideally, private access would complement public provision to maximize the health and economic gains of vaccination and minimise the potential for corruption (*34*).

Successful vaccination policies for other diseases have sometimes involved incentive payments, or penalties (*35*) or restrictions (such as refusing attendance to schools without vaccination) (*20*). Our survey shows that mandating COVID-19 vaccination commands considerable support in our global sample. Mandated COVID-19 vaccination, though, does polarize public opinion in a number of countries, the U.S. and the U.K. in particular. And there is evidence in France of overwhelming opposition. The development, and communication, of policies that in any way mandate COVID-19 vaccination should carefully consider the possible risk of creating a politically polarizing issue, as occurred with the wearing of masks in the United States.

Our survey finds that a very diverse set of allocation priorities are expressed by the general public. These encompass a concern for protecting the most vulnerable and reducing transmission while allowing life to return to normal and allowing productive sectors of the economy to open. These preferences could be used to help weight the multiple criteria on which to assess various vaccination strategies.

These results should help inform the ongoing debate over how COVID-19 vaccination programs should be implemented. In particular, they identify opportunities for policymakers, who have often struggled during the COVID-19 pandemic to meet their obligations to protect public health and the economy while simultaneously respecting the public will. While approaches to lockdowns have been divisive and politicized, a positive message from this study is that, with the exception of mandatory vaccination, the public have generally consistent preferences regarding vaccination programs and these hold across political and geographic divides.

This does not mean that government vaccination programs should fully accord with public preferences. Designing optimal vaccination programs is complex; there are many externalities to consider; and there is a clear role for expert input (*36*). Yet these programmatic choices incorporate important implicit value judgements; and the diversity of actual policies across countries emphasises the scope for experts to reach different conclusions. At a minimum, governments should take stock of public opinion. Governments, acting in the public interest, may enact vaccination programs that prioritize groups differently than would the general public. It is important though that governments recognize these differences between policy choices and public preferences and that these differences inform their efforts to gain public acceptance of their COVID-19 vaccination program.

## Data Availability

The data is not avaliable at this time as the study is subject to peer-review.

https://oxford-candour.com

## Acknowledgments

The research was supported by the National Institute for Health Research (NIHR) Oxford Biomedical Research Centre (BRC) and the COVID-19 Oxford Vaccine Trial. The views expressed are those of the author(s) and not necessarily those of the NHS, the NIHR or the Department of Health. We acknowledge the support of the Office of the Dean of the Faculty of Arts & Science at the University of Toronto. Jorge Friedman received funding from the University of Santiago project Dicyt USA1899. Alessia Melegaro acknowledges support from the Italian Ministry of Education Progetti di Rilevante Interesse Nazionale (PRIN), grant number 20177BRJXS. Jean-Francois Bonnefon acknowledges support from the grant ANR-17-EURE-0010 Investissements d’Avenir.

## Supplementary materials

### 4.1 Materials and Methods

#### Sample

The Oxford COVID-19 Vaccine Preference and Opinion Survey (CANDOUR) Project conducted online surveys of adults over 18 years of age from 13 countries. The sample of countries included Australia, Brazil, Canada, Chile, China, Colombia, France, India, Italy, Spain, Uganda, the U.S. and the U.K. They account for about half the global population and represent very diverse social and economic contexts. In each country we interviewed between 1000 and 1500 respondents during the period November 15, 2020 to December 21, 2020.

In all but Chile and Uganda, respondents were sampled by the sampling firm, Respondi.^6^ The Respondi participants were compensated for completing the survey. For these twelve countries, the modal incentive was $3.50 for a median length of interview (LOI) of 25.53 minutes. In Chile and Uganda the respondents were sampled using Facebook Ad Manager (*37*).^7^ Respondents in Chile received payments of $3.00 and in Uganda $2.25. The final sample is reasonably representative of the national population.

We implemented a quota strategy that generated samples that roughly matched the populations on age, education, gender and region. Post-stratification weights were constructed to account for remaining imbalances, as explained below. Among the panelists invited to take our survey, the response rate (calculated as the fraction of complete responses over invited, eligible participants) was 21.3%, averaged across all countries. The final sample included an average of 1,195 respondents per country (15,536 respondents overall). Descriptive statistics are reported in Table 4 and Table 5.

The survey was conducted according to the University of Oxford’s policy for human subjects research and approved by the University of Oxford Medical Sciences Interdivisional Research Ethics Committee (MS IDREC) (Approval ID: R72328/RE001). Informed consent was obtained from each participant at the beginning of the survey.

#### Experimental Design

A conjoint experiment was embedded at the beginning of the CANDOUR questionnaire on the theme of the COVID-19 vaccine. The experiment aimed to identify public preferences for which groups should be prioritised to receive limited available doses of COVID-19 vaccine. As an introduction to the overall survey, respondents were presented with a short definition of vaccines and how they work. Conjoint survey experiments are frequently employed to identify the importance individuals attribute to different features or characteristics of choices (*23*). Examples include environmental migrants (*24*), asylum seekers (*25*) and migration destinations (*26*). Awad et al. (*27*) employed conjoint experiments that generate 40 million decisions to determine the ethical principles the public thinks should guide machine behavior.^8^ We implemented a standard fully randomized paired profiles conjoint design (see Figure 4 for an example) in which each respondent was shown profiles of two different hypothetical vaccine recipients displayed side-by-side (*30*). In the case of policy-oriented survey experiments, evidence suggests that the weights given to attribute characteristics in conjoint survey experiments map closely to the actual policy choices the population would make in real world decisions, such as referendums. (*29*).

In our conjoint experiment each of the 15,536 subjects made eight binary choices over hypothetical vaccine recipients (a total of 124,288 pair-wise comparisons) that randomly varied on five attributes that are being used, or have been proposed as being important criteria for vaccine allocation.

#### Outcomes

As Figure 4 indicates, respondents were shown two potential vaccine recipients (Person A and Person B). Respondents were first asked to chose which of the potential recipients should receive the COVID-19 vaccine immediately. The resulting choice outcome variable has a value of 1 for the preferred profile and 0 for the profile that was not selected. Figure 4 has the exact wording of the question. Table 3 presents the attributes and summarizes the distribution of the randomly assigned attribute values for the global sample (these are for the English versions of the survey that were administered in Australia, Canada, India, Uganda, the U.K., and the U.S.).

#### Survey Text

The *conjoint experiment* is the first section of the survey. The experiment had a prelude introducing the vaccine allocation policy issue. Respondents were then informed of the profile attributes. Following this, respondents were asked a series of questions in which they were presented with a pair of profiles. In each question, they were asked to choose between the two profiles; and then to evaluate the extent to which their selected profile should be prioritised on a 7-point Likert scale. The wording of the items follows:

##### Prelude

Across the world, COVID-19 has infected tens of millions, killed more than one million and resulted in loss of jobs, school closures and overall economic loss. A vaccination would prevent people from getting affected by the virus. It is like the flu vaccine: Some people who get the vaccine could still get COVID-19 because it is not 100% effective; the protection could last a few months or for years; and it could have side effects. Once a reliable COVID-19 vaccine is available, health officials will give some individuals priority over others. Some individuals will get the COVID-19 vaccine immediately and other individuals will have lower priority. We are interested in your opinion about these decisions. In the next screens you will be shown two individuals (Person A and Person B) who have different characteristics that are related to COVID-19. When each screen comes up, you will be asked to choose the person (Person A or Person B) who should get the COVID-19 vaccine immediately. You will be provided with five characteristics for each individual, Person A and Person B. These five characteristics are the following: if infected, the person’s risk of COVID-19 related death compared to an average person, the person’s risk of catching and transmitting the COVID-19 virus, the income level of the person, the occupation status of the individual, and the age category of the person. We would like you to indicate which of the two persons (Person A or Person B) should have priority and receive the vaccine immediately.

##### Questions

1. *(Choice)* As you can see each of the persons (Person A and Person B) differs on our five characteristics: risk of COVID-19 related death, risk of catching and transmitting the COVID-19 virus, income level, occupation status, and age category. This vaccine could be given to one of these persons (Person A or Person B) immediately. Please indicate the person you think should get the vaccine immediately. Which of the Persons do you think should get the vaccine immediately? Select one of them. Person A; Person B.
2. *(Rating)* Person A: On a scale from 1 to 7, where 1 indicates that you think Person A should get Very Low Priority for the vaccine and 7 indicates that you think Person A should get Very High Priority for the vaccine, what vaccine priority would you give Person A?

The survey instrument included a number of different themes in addition to the conjoint experiment. Many of the variables are described in Table 4 and Table 5. A full version of the survey instrument is included in these Supplementary Materials. Here we present the wording of the questions presented in Figure 3:

- Talking about vaccines in general, in some countries vaccines are only available from the government either at low or no cost. In some countries vaccines are only available for private purchase. And in some countries vaccines are available from the government but citizens can pay privately to gain early access. Which of these three approaches do you think should be applied to the COVID-19 vaccine? Would you prefer
  – Vaccines only made available by government at low or no cost?
  – Vaccines are only available for private purchase?
  – Vaccines made available by government but citizens can pay privately to gain access?
- Consider the following situation: a COVID-19 vaccine becomes available and is provided by government health agencies. For 80 out of 100 people the vaccine would provide protection for at least 18 months. But there are limited initial supplies of the vaccine. For this reason, you would have to wait 6 months before you could receive it. If a COVID-19 vaccine was also available for private purchase and you could receive it immediately would you considering buying it?
  – Yes
  – No
  – Do not know
- We are interested in your opinion about the implementation of the COVID-19 vaccine once it is available. Please use the sliding scale to indicate how much you agree or disagree with the statements. You can move the pointer from 0 which means very much disagree to 100 which means very much agree.
  – The government should make the COVID-19 vaccine mandatory for everybody.

#### Survey translations

The survey was translated from English into five languages: Chinese, French, Italian, Spanish and Portuguese. These translations were conducted by professional translators. They were then reviewed by a number of native speakers. They were then translated back to English.

#### Statistical analysis

The 15,536 respondents from the 13-country sample viewed eight pairs of vaccine allocation profiles and made a forced choice (i.e. their was no opt-out option). This generated 248,576 profile evaluations. Our goal in the multivariable modeling of these dichotomous outcomes was simply to identify which of the attribute values were more (or less) likely to cause respondents to select a particular profile. The logistic models regressed the forced vaccine allocation priority choices on sets of indicator variables that represent the values of each attribute while omitting one level of each attribute as the reference category and clustering the standard errors by respondent. Given that the attribute values were randomly assigned across all profiles and respondents these are reasonable conclusions to draw from the logistic coefficients. These logistic coefficients, along with their standard errors clustered at respondent level, are reported in the main text. In these Supplementary Tables we also report the average marginal component-specific effects (AMCEs) of each attribute on respondents’ choice of a vaccine allocation priority, which we were able to identify due to the random assignment of attribute values across all profiles and respondents (*30*). For the AMCEs we estimated identical models to those we estimated with logistic regression – again, regressing the forced vaccine allocation priority choice on the same sets of indicator variables. For the AMCEs we employed linear least squares regression, again with standard errors clustered by respondent. The linear least squares regression results generated substantively identical results to those from the logistic regression reported in the manuscript.

### 4.2 Supplementary Text

#### Notes on conjoint plots and regression tables

1. All plots of the logistic regression coefficients and the linear regression AMCEs contained in figures and in the this Supplementary Material document are based on logistic regression and OLS linear regression. The dots with horizontal lines indicate point estimates with cluster-robust 95% confidence intervals. The unfilled dots on the zero line denote reference categories for each vaccine allocation attribute.
2. The estimated logistic coefficient and AMCEs displayed in plots are numerically presented in corresponding regression tables in this Supplementary Materials document. Captions to all relevant figures include a reference to a corresponding regression table.
3. All regression models were estimated with logistic regression or OLS linear regression. Point estimates are displayed and standard errors clustered by respondent are shown in parentheses in the regression tables.
4. Unless otherwise noted, all regression models employed the choice outcome as the dependent variable (forced choice).
5. Unless otherwise noted, all regression models regressed the dependent variable on the full set of vaccine attribution attributes (a dummy variable for all levels except for one reference level per attribute).
6. For all regression models, the reference categories for the five attributes were as follows: Age: 25 years old, Income: Lowest 20% income level, Occupation: Not working, Transmission risk: Average risk of transmission, Risk of death: Average risk of death.
7. In each model, the number of observations reflects the number of survey respondents pertaining to that model multiplied by 16, since each respondent evaluated 16 profiles (8 pairs) in total.

### 4.3 Supplementary Figures and Tables

**Figure 4:**
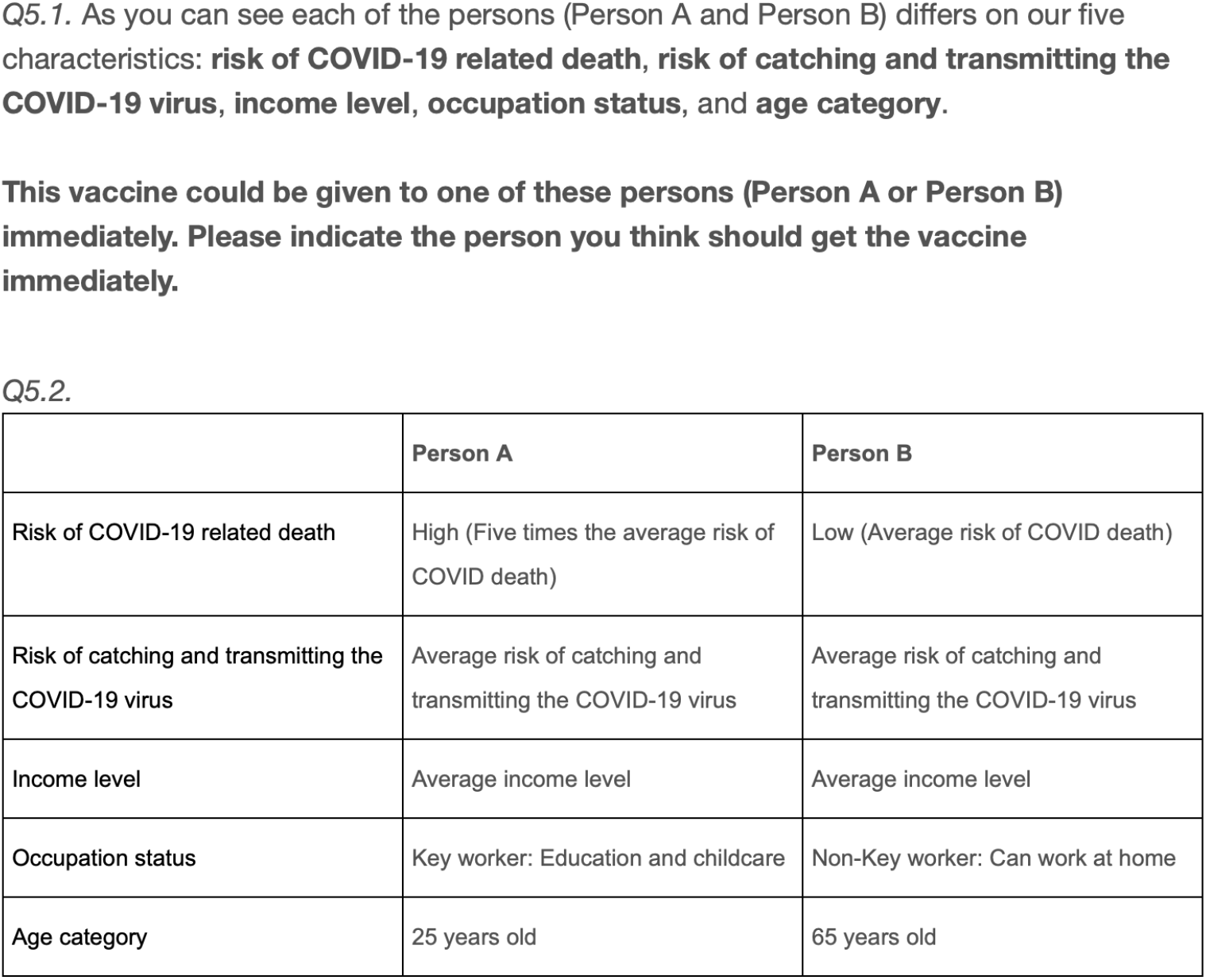
Example profile pair. This figure displays an example of a pair of vaccine recipient profiles seen by the survey respondents. This example comes from the English version of the survey administered to respondents in the United Kingdom. Each respondent saw and evaluated eight separate pairs. The order of the attributes (rows) was fully randomized between respondents, but for each respondent, the order was kept constant across the five pairs they were shown. The specific attribute levels (values in the cells in the last two columns) were fully randomized between and within respondents.

**Figure 5:**
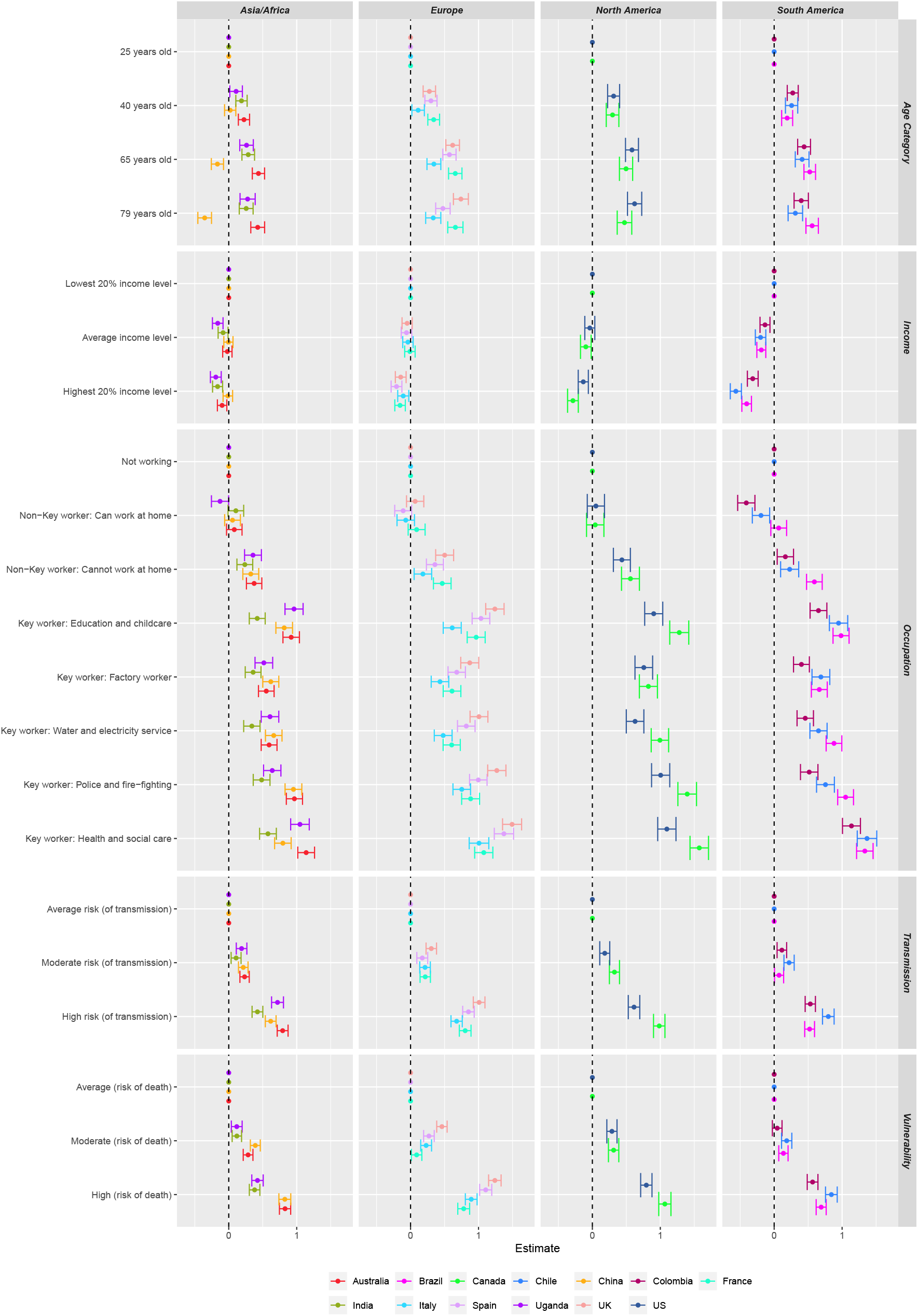
Vaccine Candidate Decisions: Logistic coefficients are reported for attribute values. 95% confidence intervals are shown for each point estimate, clustered by subject.

**Figure 6:**
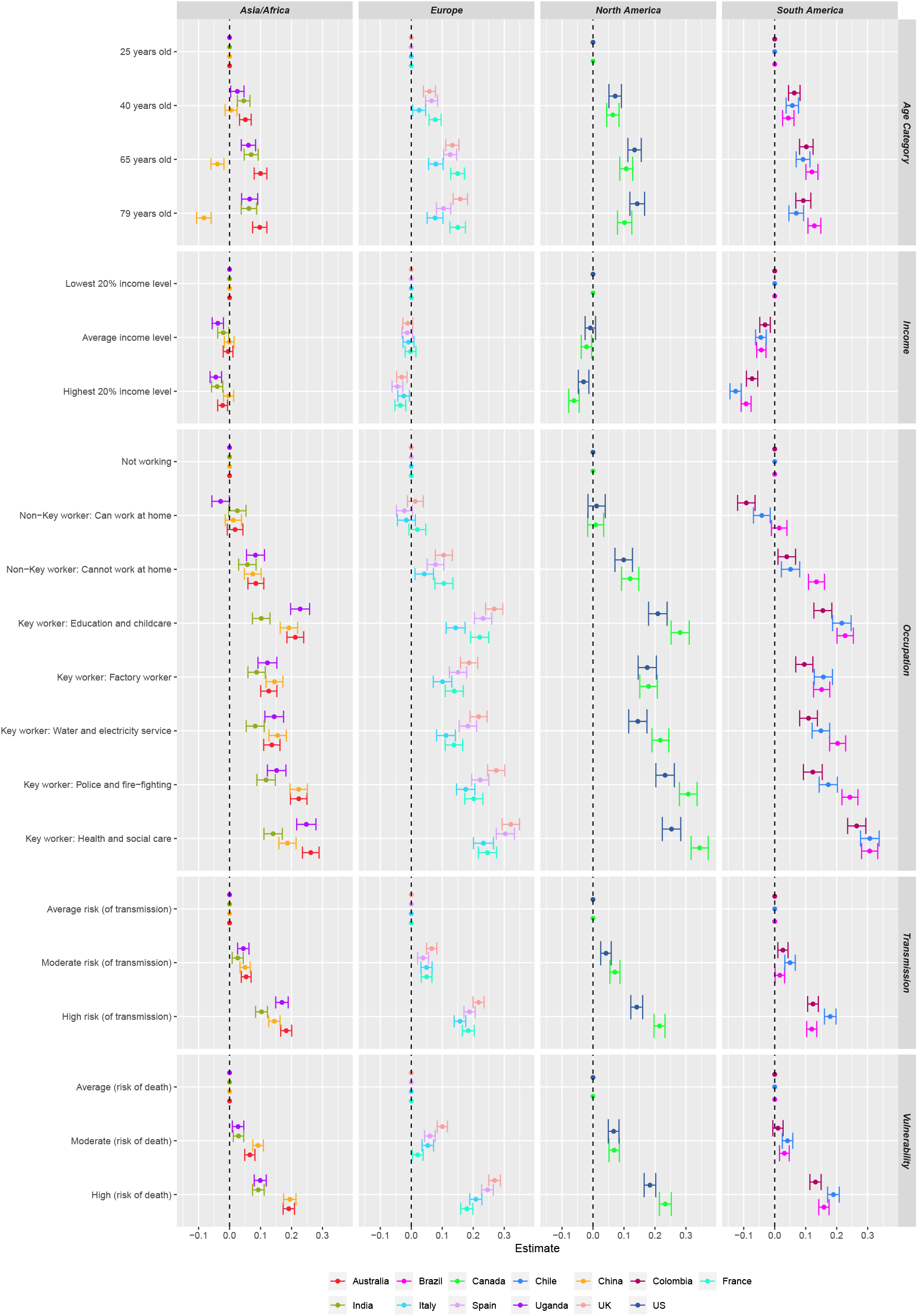
Vaccine Candidate Decisions: Linear regression coefficients are reported for attribute values. 95% confidence intervals are shown for each point estimate, clustered by subject.

**Figure 7:**
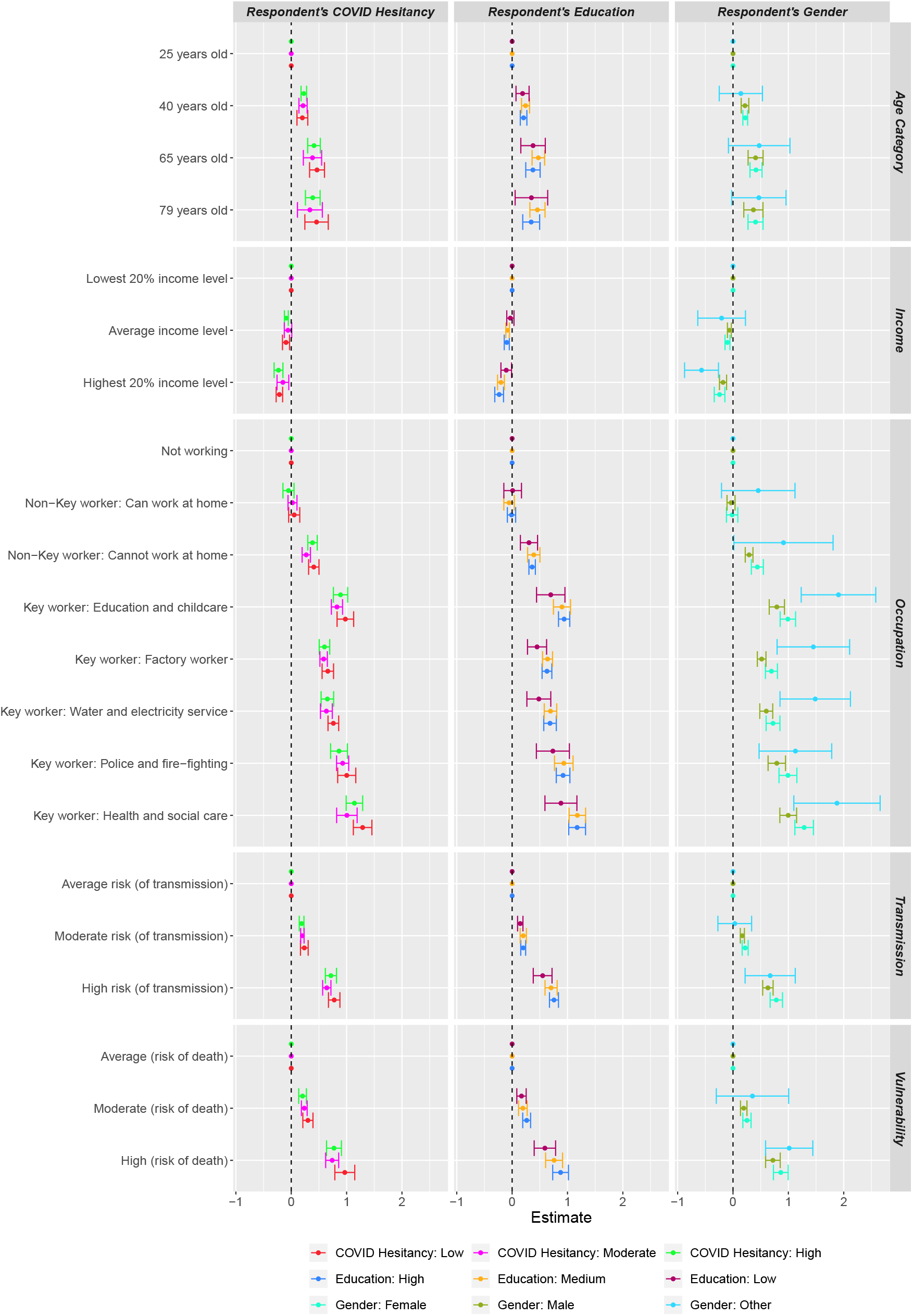
Vaccine Candidate Decisions by COVID-19 Vaccine Hesitancy, Education and Gender: Logistic coefficients are reported for attribute values. 95% confidence intervals are shown for each point estimate, clustered by subject.

**Figure 8:**
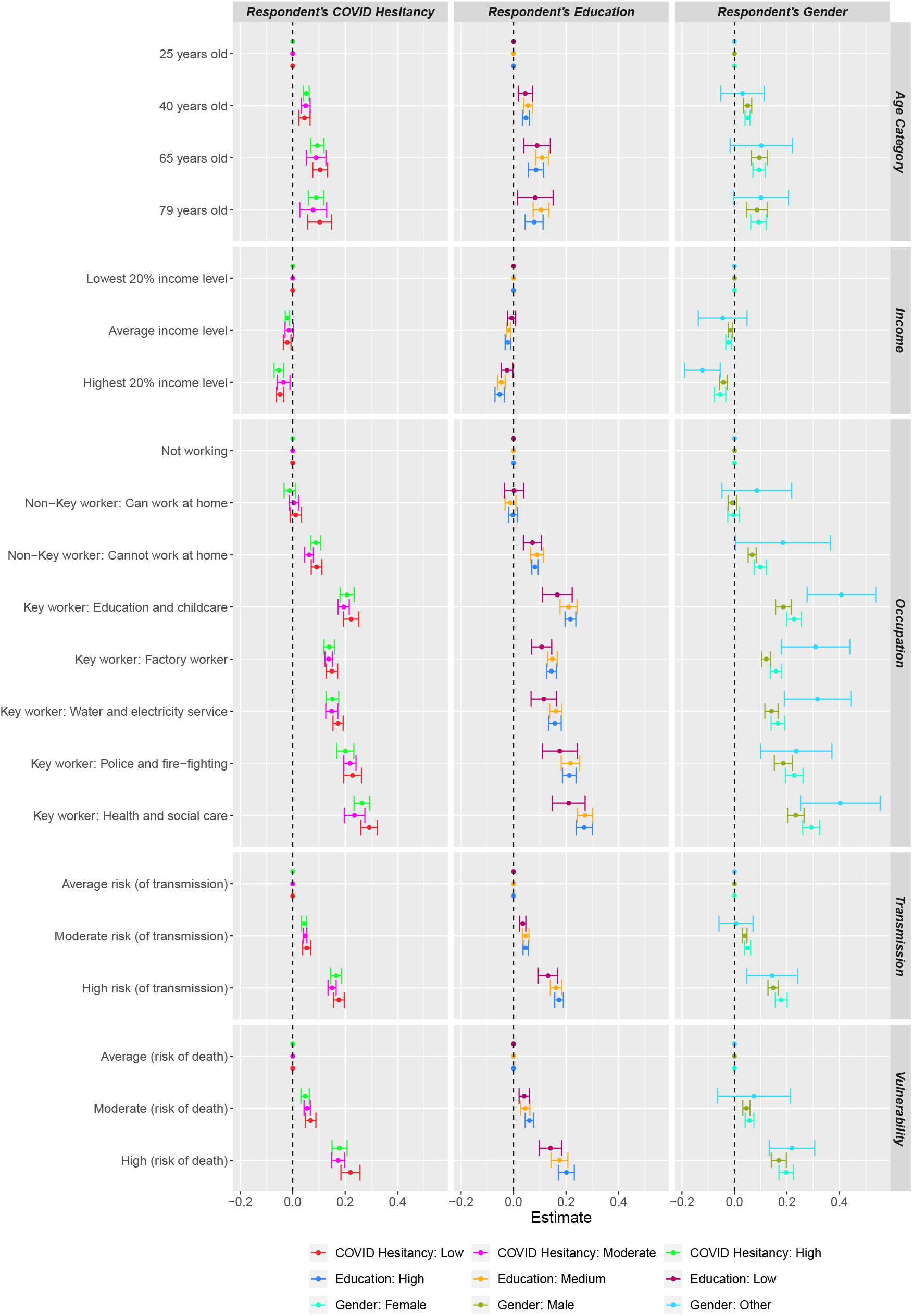
Vaccine Candidate Decisions by COVID-19 Vaccine Hesitancy, Education and Gender: Linear regression coefficients are reported for attribute values. 95% confidence intervals are shown for each point estimate, clustered by subject.

**Figure 9:**
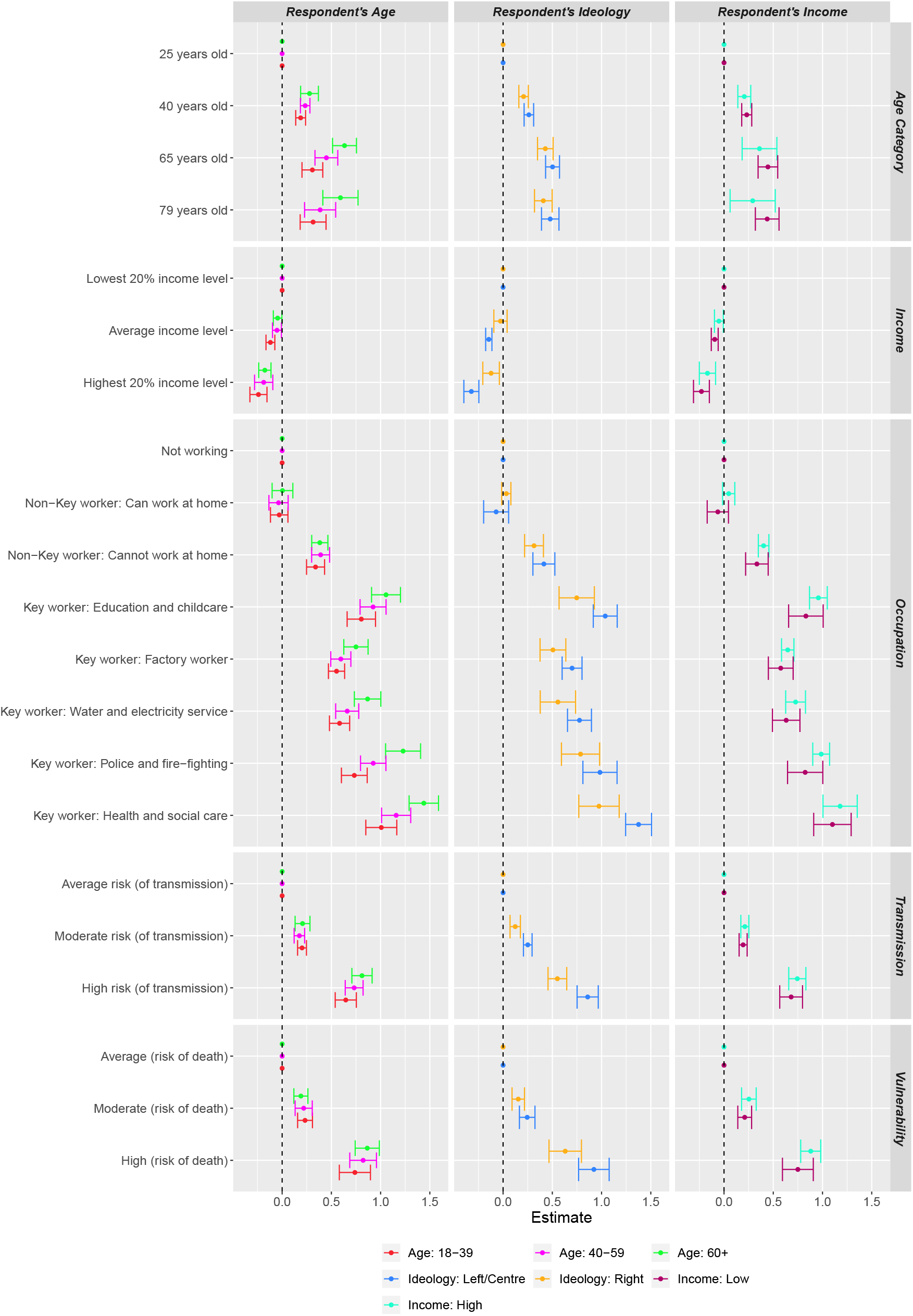
Vaccine Age, Left-Right Identification and Income: Logistic regression coefficients are reported for attribute values. 95% confidence intervals are shown for each point estimate, clustered by subject.

**Figure 10:**
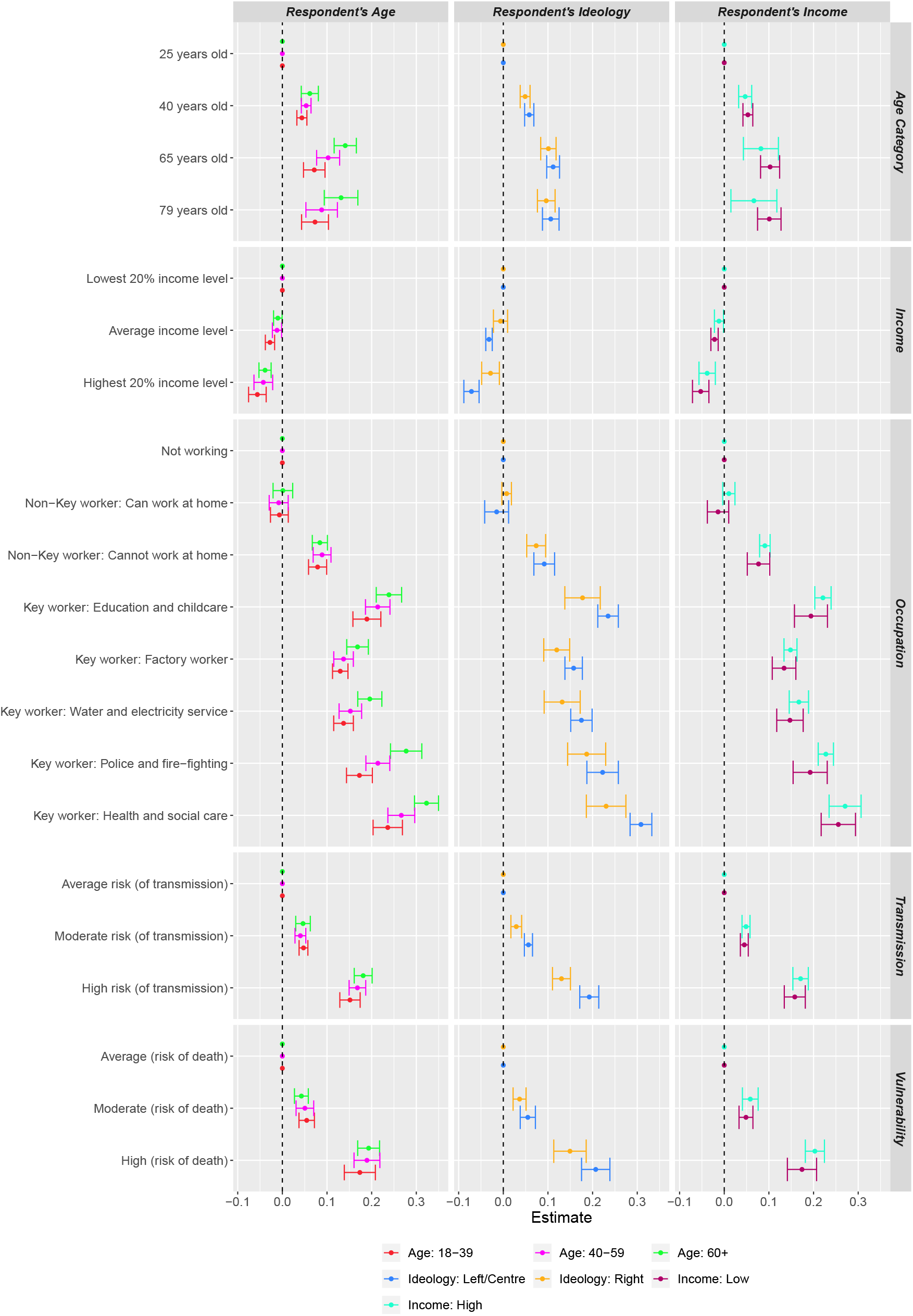
Vaccine Age, Left-Right Identification and Income: Linear regression coefficients are reported for attribute values. 95% confidence intervals are shown for each point estimate, clustered by subject.

**Table 2:** Global Sample Details

|  | Sample Size | Recruit | Language | Begin Date | End Date |
| --- | --- | --- | --- | --- | --- |
| Australia | 1364 | Respondi | English | 27/11/2020 | 24/12/2020 |
| Brazil | 1426 | Respondi | Portuguese | 27/11/2020 | 24/12/2020 |
| Canada | 1150 | Respondi | English/French | 27/11/2020 | 24/12/2020 |
| Chile | 1122 | Santiago CESS | Spanish | 27/11/2020 | 13/12/2020 |
| China | 1298 | Respondi | Chinese | 24/11/2020 | 24/12/2020 |
| Colombia | 1237 | Respondi | Spanish | 27/11/2020 | 28/12/2020 |
| France | 1146 | Respondi | French | 27/11/2020 | 28/12/2020 |
| India | 1191 | Respondi | English | 17/12/2020 | 5/01/2021 |
| Italy | 1081 | Respondi | Italian | 27/11/2020 | 28/12/2020 |
| Spain | 1153 | Respondi | Spanish | 27/11/2020 | 24/12/2020 |
| Uganda | 1053 | Facebook | English | 17/12/2020 | 14/01/2021 |
| UK | 1165 | Respondi | English | 27/11/2020 | 28/12/2020 |
| US | 1150 | Respondi | English | 27/11/2020 | 24/12/2020 |

**Table 3:**
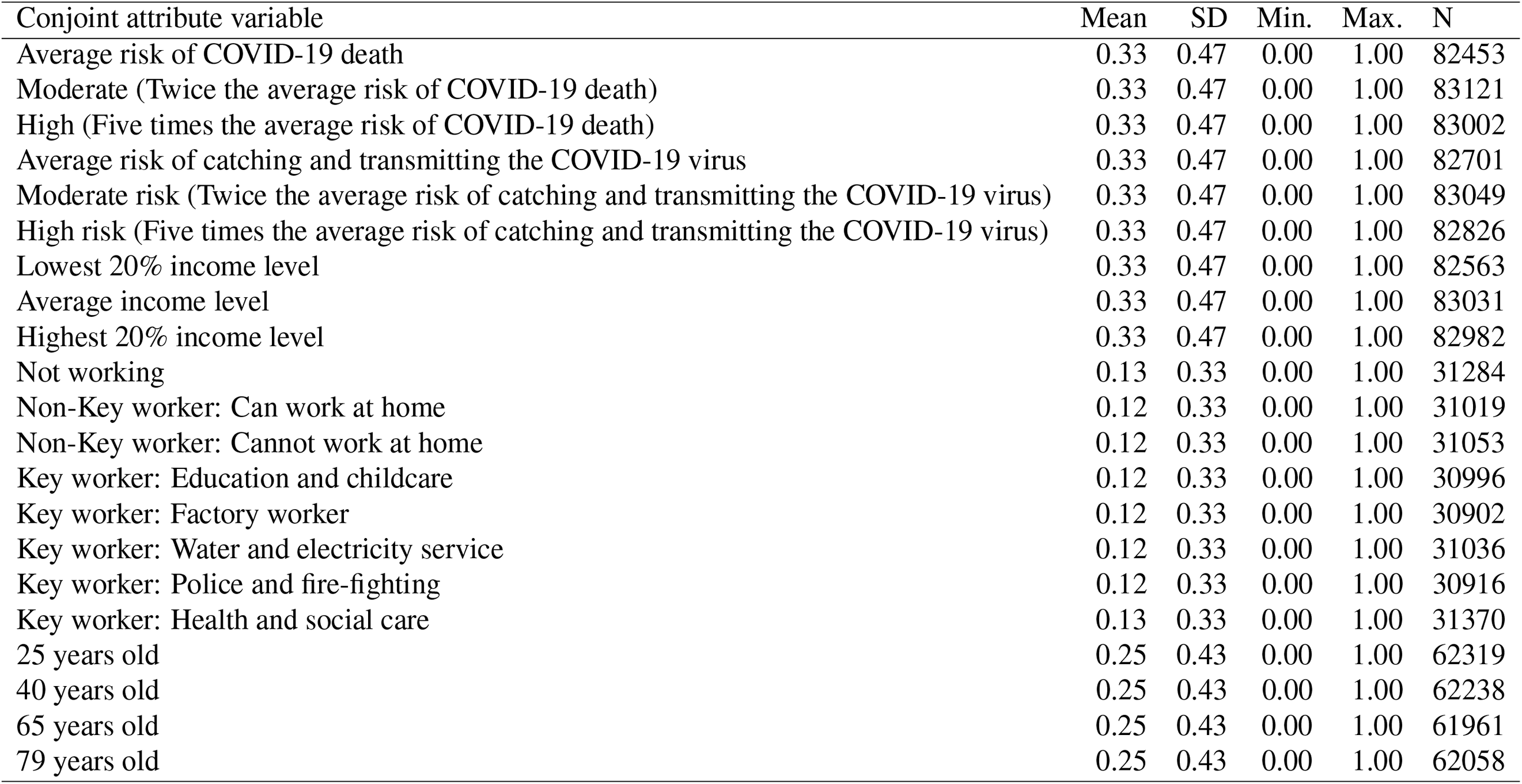
Combined Global Survey: Summary of Conjoint Attribute Randomisation

**Table 4:** Summary statistics. This table shows the proportion of subjects within each country along key demographics.

|  | **Age** |  |  | **Ideology** |  | **Income** |  |
| --- | --- | --- | --- | --- | --- | --- | --- |
|  | *18-39* | *40-59* | *60+* | *Left/Centre* | *Right* | *Low* | *High* |
| Australia | 42.4% | 33.8% | 23.9% | 55.1% | 44.9% | 59.5% | 40.5% |
| Brazil | 48.2% | 35.9% | 15.8% | 50.9% | 49.1% | 52.1% | 47.9% |
| Canada | 38.9% | 33.7% | 27.4% | 68% | 32% | 55.4% | 44.6% |
| Chile | 60.1% | 31.6% | 8.3% | 80.4% | 19.6% | 73.7% | 26.3% |
| China | 55.9% | 34.4% | 9.6% | 0% | 0% | 19.1% | 80.9% |
| Colombia | 54.4% | 38.2% | 7.4% | 68.9% | 31.1% | 64% | 36% |
| France | 29.3% | 35.6% | 35.1% | 62.7% | 37.3% | 67.6% | 32.4% |
| India | 78.6% | 17.1% | 4.3% | 27.7% | 72.3% | 81.6% | 18.4% |
| Italy | 32.4% | 42.1% | 25.5% | 54.5% | 45.5% | 56.6% | 43.4% |
| Spain | 32.6% | 39.4% | 28% | 73.1% | 26.9% | 56.4% | 43.6% |
| Uganda | 91.3% | 8.4% | 0.4% | 55.7% | 44.3% | 96.7% | 3.3% |
| UK | 29.3% | 36.9% | 33.8% | 61.2% | 38.8% | 50.3% | 49.7% |
| US | 36.2% | 36.5% | 27.3% | 50.4% | 49.6% | 47.6% | 52.4% |

**Table 5:** Summary statistics, continued. This table shows the proportion of subjects within each country along key demographics.

|  | **COVID-19 Vaccine Hesitancy** |  |  | **Education** |  |  | **Gender** |  |  |
| --- | --- | --- | --- | --- | --- | --- | --- | --- | --- |
|  | *Low* | *Moderate* | *High* | *Low* | *Medium* | *High* | *Female* | *Male* | *Other* |
| Australia | 18.3% | 21.1% | 60.6% | 30.7% | 26.5% | 42.8% | 52.5% | 47.4% | 0.1% |
| Brazil | 11% | 16.8% | 72.2% | 19.1% | 32.3% | 48.6% | 50.2% | 49.7% | 0.1% |
| Canada | 20% | 20.2% | 59.8% | 5.1% | 48.2% | 46.7% | 46% | 53.7% | 0.3% |
| Chile | 10.1% | 10.5% | 79.4% | 1.1% | 49.1% | 49.8% | 60.6% | 38.9% | 0.4% |
| China | 14.7% | 34.4% | 50.9% | 24% | 15.9% | 60.1% | 47.1% | 52.8% | 0.1% |
| Colombia | 7.6% | 8.7% | 83.7% | 15.2% | 42.6% | 42.3% | 57.6% | 42.2% | 0.2% |
| France | 9.3% | 13.6% | 77.1% | 3.7% | 55.7% | 40.5% | 44.4% | 55.6% | 0% |
| India | 10.4% | 16.2% | 73.5% | 19.9% | 30.3% | 49.8% | 39.3% | 60.5% | 0.2% |
| Italy | 21.1% | 27.8% | 51.1% | 23.6% | 49% | 27.4% | 54.6% | 45.2% | 0.2% |
| Spain | 8.1% | 11.6% | 80.3% | 9.6% | 23.6% | 66.8% | 51.3% | 48.6% | 0.1% |
| Uganda | 8.6% | 10.8% | 80.5% | 0% | 36.3% | 63.7% | 25.5% | 73.4% | 1.1% |
| UK | 29.1% | 21.8% | 49.1% | 22.9% | 42.4% | 34.6% | 46.1% | 53.7% | 0.2% |
| US | 13.2% | 18.6% | 68.3% | 3.9% | 46% | 50.2% | 49.3% | 50.6% | 0.1% |

**Table 6:** Country Logistic Regression Results. The dependent variable is the Forced Choice decision. These are the estimates used to construct the conjoint plots presented in Figure 1.

|  | Country Model |  |  |  |  |  |  |  |  |  |  |  |  |
| --- | --- | --- | --- | --- | --- | --- | --- | --- | --- | --- | --- | --- | --- |
|  | Canada | Colombia | Spain | US | Australia | UK | France | Italy | China | Brazil | Chile | Uganda | India |
| (Intercept) | -1.92***<br>(0.07) | -0.91***<br>(0.06) | -1.68***<br>(0.07) | -1.57***<br>(0.07) | -1.52***<br>(0.07) | -2.15***<br>(0.07) | -1.58***<br>(0.07) | -1.24***<br>(0.07) | -1.08***<br>(0.07) | -1.29***<br>(0.06) | -1.23***<br>(0.07) | -1.03***<br>(0.07) | -0.76***<br>(0.06) |
| Vulnerability |  |  |  |  |  |  |  |  |  |  |  |  |  |
| Moderate (risk of death) | 0.31***<br>(0.04) | 0.04<br>(0.04) | 0.27***<br>(0.04) | 0.29***<br>(0.04) | 0.28***<br>(0.04) | 0.46***<br>(0.04) | 0.09*<br>(0.04) | 0.23***<br>(0.04) | 0.39***<br>(0.04) | 0.14***<br>(0.03) | 0.18***<br>(0.04) | 0.12**<br>(0.04) | 0.12***<br>(0.04) |
| High (risk of death) | 1.07***<br>(0.05) | 0.57***<br>(0.04) | 1.11***<br>(0.05) | 0.79***<br>(0.04) | 0.83***<br>(0.04) | 1.24***<br>(0.05) | 0.78***<br>(0.05) | 0.89***<br>(0.04) | 0.83***<br>(0.04) | 0.69***<br>(0.04) | 0.84***<br>(0.04) | 0.42***<br>(0.04) | 0.38***<br>(0.04) |
| Transmission |  |  |  |  |  |  |  |  |  |  |  |  |  |
| Moderate risk (of transmission) | 0.33***<br>(0.04) | 0.11**<br>(0.04) | 0.17***<br>(0.04) | 0.18***<br>(0.04) | 0.23***<br>(0.04) | 0.31***<br>(0.04) | 0.22***<br>(0.04) | 0.21***<br>(0.04) | 0.21***<br>(0.04) | 0.07*<br>(0.03) | 0.22***<br>(0.04) | 0.19***<br>(0.04) | 0.11***<br>(0.04) |
| High risk (of transmission) | 0.98***<br>(0.04) | 0.53***<br>(0.04) | 0.85***<br>(0.04) | 0.61***<br>(0.04) | 0.79***<br>(0.04) | 1.01***<br>(0.04) | 0.80***<br>(0.04) | 0.68***<br>(0.04) | 0.62***<br>(0.04) | 0.52***<br>(0.04) | 0.80***<br>(0.04) | 0.72***<br>(0.04) | 0.42***<br>(0.04) |
| Income |  |  |  |  |  |  |  |  |  |  |  |  |  |
| Average income level | -0.10*<br>(0.04) | -0.13***<br>(0.04) | -0.06<br>(0.04) | -0.04<br>(0.04) | -0.02<br>(0.04) | -0.05<br>(0.04) | -0.01<br>(0.04) | -0.04<br>(0.04) | -0.01<br>(0.03) | -0.19***<br>(0.03) | -0.20***<br>(0.04) | -0.16***<br>(0.04) | -0.08*<br>(0.04) |
| Highest 20% income level | -0.29***<br>(0.04) | -0.32***<br>(0.04) | -0.21***<br>(0.04) | -0.13***<br>(0.04) | -0.10**<br>(0.04) | -0.15***<br>(0.04) | -0.15***<br>(0.04) | -0.11**<br>(0.04) | -0.01<br>(0.04) | -0.40***<br>(0.04) | -0.56***<br>(0.04) | -0.19***<br>(0.04) | -0.16***<br>(0.04) |
| Occupation |  |  |  |  |  |  |  |  |  |  |  |  |  |
| Non-Key worker: Can work at home | 0.04<br>(0.07) | -0.41***<br>(0.06) | -0.11<br>(0.06) | 0.05<br>(0.06) | 0.08<br>(0.06) | 0.07<br>(0.06) | 0.09<br>(0.06) | -0.07<br>(0.07) | 0.06<br>(0.06) | 0.07<br>(0.06) | -0.19**<br>(0.07) | -0.13*<br>(0.06) | 0.10<br>(0.06) |
| Non-Key worker: Cannot work at home | 0.56***<br>(0.07) | 0.16***<br>(0.06) | 0.36***<br>(0.06) | 0.43***<br>(0.06) | 0.37***<br>(0.06) | 0.50***<br>(0.07) | 0.47***<br>(0.07) | 0.18***<br>(0.07) | 0.32***<br>(0.06) | 0.59***<br>(0.06) | 0.23***<br>(0.07) | 0.36***<br>(0.06) | 0.24***<br>(0.06) |
| Key worker: Education and childcare | 1.28***<br>(0.07) | 0.65***<br>(0.06) | 1.04***<br>(0.07) | 0.90***<br>(0.07) | 0.92***<br>(0.06) | 1.24***<br>(0.07) | 0.97***<br>(0.07) | 0.61***<br>(0.07) | 0.82***<br>(0.06) | 0.98***<br>(0.06) | 0.95***<br>(0.07) | 0.96***<br>(0.07) | 0.42***<br>(0.06) |
| Key worker: Factory worker | 0.82***<br>(0.07) | 0.40***<br>(0.06) | 0.68***<br>(0.07) | 0.76***<br>(0.07) | 0.55***<br>(0.06) | 0.87***<br>(0.07) | 0.61***<br>(0.07) | 0.43***<br>(0.07) | 0.62***<br>(0.06) | 0.66***<br>(0.06) | 0.69***<br>(0.07) | 0.52***<br>(0.07) | 0.36***<br>(0.06) |
| Key worker: Water and electricity service | 0.99***<br>(0.07) | 0.46***<br>(0.06) | 0.82***<br>(0.07) | 0.63***<br>(0.07) | 0.59***<br>(0.06) | 1.01***<br>(0.07) | 0.61***<br>(0.07) | 0.48***<br>(0.07) | 0.66***<br>(0.06) | 0.88***<br>(0.06) | 0.65***<br>(0.06) | 0.61***<br>(0.07) | 0.34***<br>(0.06) |
| Key worker: Police and fire-fighting | 1.40***<br>(0.07) | 0.52***<br>(0.07) | 1.00***<br>(0.07) | 1.01***<br>(0.07) | 0.97***<br>(0.06) | 1.27***<br>(0.07) | 0.88***<br>(0.07) | 0.75***<br>(0.07) | 0.95***<br>(0.06) | 1.05***<br>(0.06) | 0.76***<br>(0.07) | 0.64***<br>(0.07) | 0.48***<br>(0.06) |
| Key worker: Health and social care | 1.57***<br>(0.07) | 1.14***<br>(0.07) | 1.37***<br>(0.07) | 1.10***<br>(0.07) | 1.14***<br>(0.06) | 1.49***<br>(0.07) | 1.08***<br>(0.07) | 1.01***<br>(0.07) | 0.80***<br>(0.06) | 1.33***<br>(0.06) | 1.37***<br>(0.07) | 1.05***<br>(0.07) | 0.58***<br>(0.06) |
| Age Category |  |  |  |  |  |  |  |  |  |  |  |  |  |
| 40 years old | 0.30***<br>(0.05) | 0.27***<br>(0.04) | 0.30***<br>(0.05) | 0.31***<br>(0.04) | 0.22***<br>(0.04) | 0.28***<br>(0.05) | 0.34***<br>(0.05) | 0.11*<br>(0.05) | 0.02<br>(0.04) | 0.19***<br>(0.04) | 0.26***<br>(0.05) | 0.11*<br>(0.05) | 0.19***<br>(0.04) |
| 65 years old | 0.50***<br>(0.05) | 0.44***<br>(0.05) | 0.57***<br>(0.05) | 0.58***<br>(0.05) | 0.44***<br>(0.05) | 0.62***<br>(0.05) | 0.66***<br>(0.05) | 0.34***<br>(0.05) | -0.17***<br>(0.05) | 0.52***<br>(0.04) | 0.41***<br>(0.05) | 0.26***<br>(0.05) | 0.29***<br>(0.05) |
| 79 years old | 0.47***<br>(0.05) | 0.40***<br>(0.05) | 0.48***<br>(0.05) | 0.62***<br>(0.05) | 0.43***<br>(0.05) | 0.74***<br>(0.06) | 0.66***<br>(0.06) | 0.33***<br>(0.06) | -0.35***<br>(0.05) | 0.56***<br>(0.05) | 0.31***<br>(0.05) | 0.28***<br>(0.06) | 0.26***<br>(0.05) |

\*\*\*p < 0.001; \*\*p < 0.01; \*p < 0.05
\*\*\* $p < 0.001$ ; \*\* $p < 0.01$ ; \* $p < 0.05$

**Table 7:** Country Linear (OLS) Regression Results. The dependent variable is the Forced Choice decision. These are the estimates used to construct the conjoint plots presented in Figure 6.

|  | Country Model |  |  |  |  |  |  |  |  |  |  |  |  |
| --- | --- | --- | --- | --- | --- | --- | --- | --- | --- | --- | --- | --- | --- |
|  | Canada | Colombia | Spain | US | Australia | UK | France | Italy | China | Brazil | Chile | Uganda | India |
| (Intercept) | 0.08***<br>(0.01) | 0.29***<br>(0.01) | 0.13***<br>(0.01) | 0.14***<br>(0.01) | 0.15***<br>(0.01) | 0.04***<br>(0.01) | 0.14***<br>(0.01) | 0.21***<br>(0.02) | 0.25***<br>(0.02) | 0.20***<br>(0.01) | 0.22***<br>(0.02) | 0.26***<br>(0.02) | 0.31***<br>(0.02) |
| Vulnerability |  |  |  |  |  |  |  |  |  |  |  |  |  |
| Moderate (risk of death) | 0.07***<br>(0.01) | 0.01<br>(0.01) | 0.06***<br>(0.01) | 0.07***<br>(0.01) | 0.07***<br>(0.01) | 0.10***<br>(0.01) | 0.02*<br>(0.01) | 0.05***<br>(0.01) | 0.09***<br>(0.01) | 0.03***<br>(0.01) | 0.04***<br>(0.01) | 0.03***<br>(0.01) | 0.03***<br>(0.01) |
| High (risk of death) | 0.23***<br>(0.01) | 0.13***<br>(0.01) | 0.25***<br>(0.01) | 0.18***<br>(0.01) | 0.19***<br>(0.01) | 0.27***<br>(0.01) | 0.18***<br>(0.01) | 0.21***<br>(0.01) | 0.19***<br>(0.01) | 0.16***<br>(0.01) | 0.19***<br>(0.01) | 0.10***<br>(0.01) | 0.09***<br>(0.01) |
| Transmission |  |  |  |  |  |  |  |  |  |  |  |  |  |
| Moderate risk (of transmission) | 0.07***<br>(0.01) | 0.03**<br>(0.01) | 0.04***<br>(0.01) | 0.04***<br>(0.01) | 0.05***<br>(0.01) | 0.07***<br>(0.01) | 0.05***<br>(0.01) | 0.05***<br>(0.01) | 0.05***<br>(0.01) | 0.02*<br>(0.01) | 0.05***<br>(0.01) | 0.04***<br>(0.01) | 0.03***<br>(0.01) |
| High risk (of transmission) | 0.21***<br>(0.01) | 0.12***<br>(0.01) | 0.19***<br>(0.01) | 0.14***<br>(0.01) | 0.18***<br>(0.01) | 0.22***<br>(0.01) | 0.18***<br>(0.01) | 0.16***<br>(0.01) | 0.14***<br>(0.01) | 0.12***<br>(0.01) | 0.18***<br>(0.01) | 0.17***<br>(0.01) | 0.10***<br>(0.01) |
| Income |  |  |  |  |  |  |  |  |  |  |  |  |  |
| Average income level | -0.02*<br>(0.01) | -0.03***<br>(0.01) | -0.01<br>(0.01) | -0.01<br>(0.01) | -0.00<br>(0.01) | -0.01<br>(0.01) | -0.00<br>(0.01) | -0.01<br>(0.01) | -0.00<br>(0.01) | -0.04***<br>(0.01) | -0.04***<br>(0.01) | -0.04***<br>(0.01) | -0.02*<br>(0.01) |
| Highest 20% income level | -0.06***<br>(0.01) | -0.07***<br>(0.01) | -0.04***<br>(0.01) | -0.03***<br>(0.01) | -0.02***<br>(0.01) | -0.03***<br>(0.01) | -0.04***<br>(0.01) | -0.02***<br>(0.01) | -0.00<br>(0.01) | -0.09***<br>(0.01) | -0.13***<br>(0.01) | -0.04***<br>(0.01) | -0.04***<br>(0.01) |
| Occupation |  |  |  |  |  |  |  |  |  |  |  |  |  |
| Non-Key worker: Can work at home | 0.01<br>(0.01) | -0.09***<br>(0.01) | -0.02<br>(0.01) | 0.01<br>(0.01) | 0.02<br>(0.01) | 0.01<br>(0.01) | 0.02<br>(0.01) | -0.02<br>(0.01) | 0.01<br>(0.01) | 0.01<br>(0.01) | -0.04**<br>(0.01) | -0.03*<br>(0.01) | 0.03<br>(0.01) |
| Non-Key worker: Cannot work at home | 0.12***<br>(0.01) | 0.04***<br>(0.01) | 0.08***<br>(0.01) | 0.10***<br>(0.01) | 0.08***<br>(0.01) | 0.11***<br>(0.01) | 0.11***<br>(0.01) | 0.04***<br>(0.02) | 0.07***<br>(0.01) | 0.13***<br>(0.01) | 0.05***<br>(0.02) | 0.08***<br>(0.01) | 0.06***<br>(0.01) |
| Key worker: Education and childcare | 0.28***<br>(0.01) | 0.16***<br>(0.01) | 0.23***<br>(0.01) | 0.21***<br>(0.02) | 0.21***<br>(0.01) | 0.27***<br>(0.01) | 0.22***<br>(0.01) | 0.14***<br>(0.02) | 0.19***<br>(0.01) | 0.23***<br>(0.01) | 0.22***<br>(0.02) | 0.23***<br>(0.02) | 0.10***<br>(0.01) |
| Key worker: Factory worker | 0.18***<br>(0.01) | 0.10***<br>(0.01) | 0.15***<br>(0.01) | 0.18***<br>(0.01) | 0.13***<br>(0.01) | 0.19***<br>(0.01) | 0.14***<br>(0.01) | 0.10***<br>(0.02) | 0.14***<br>(0.01) | 0.15***<br>(0.01) | 0.16***<br>(0.02) | 0.12***<br>(0.02) | 0.09***<br>(0.01) |
| Key worker: Water and electricity service | 0.22***<br>(0.01) | 0.11***<br>(0.01) | 0.18***<br>(0.01) | 0.14***<br>(0.02) | 0.14***<br>(0.01) | 0.22***<br>(0.01) | 0.14***<br>(0.01) | 0.11***<br>(0.02) | 0.16***<br>(0.01) | 0.20***<br>(0.01) | 0.15***<br>(0.01) | 0.14***<br>(0.02) | 0.08***<br>(0.01) |
| Key worker: Police and fire-fighting | 0.31***<br>(0.01) | 0.12***<br>(0.02) | 0.22***<br>(0.01) | 0.23***<br>(0.02) | 0.22***<br>(0.01) | 0.27***<br>(0.01) | 0.20***<br>(0.01) | 0.18***<br>(0.02) | 0.22***<br>(0.01) | 0.24***<br>(0.01) | 0.17***<br>(0.01) | 0.15***<br>(0.02) | 0.12***<br>(0.01) |
| Key worker: Health and social care | 0.34***<br>(0.01) | 0.26***<br>(0.01) | 0.30***<br>(0.01) | 0.25***<br>(0.02) | 0.26***<br>(0.01) | 0.32***<br>(0.01) | 0.25***<br>(0.01) | 0.23***<br>(0.02) | 0.19***<br>(0.01) | 0.31***<br>(0.01) | 0.31***<br>(0.02) | 0.25***<br>(0.02) | 0.14***<br>(0.02) |
| Age Category |  |  |  |  |  |  |  |  |  |  |  |  |  |
| 40 years old | 0.06***<br>(0.01) | 0.06***<br>(0.01) | 0.07***<br>(0.01) | 0.07***<br>(0.01) | 0.05***<br>(0.01) | 0.06***<br>(0.01) | 0.08***<br>(0.01) | 0.03*<br>(0.01) | 0.00<br>(0.01) | 0.04***<br>(0.01) | 0.06***<br>(0.01) | 0.03*<br>(0.01) | 0.05***<br>(0.01) |
| 65 years old | 0.11***<br>(0.01) | 0.10***<br>(0.01) | 0.13***<br>(0.01) | 0.13***<br>(0.01) | 0.10***<br>(0.01) | 0.13***<br>(0.01) | 0.15***<br>(0.01) | 0.08***<br>(0.01) | -0.04***<br>(0.01) | 0.12***<br>(0.01) | 0.09***<br>(0.01) | 0.06***<br>(0.01) | 0.07***<br>(0.01) |
| 79 years old | 0.10***<br>(0.01) | 0.09***<br>(0.01) | 0.10***<br>(0.01) | 0.14***<br>(0.01) | 0.10***<br>(0.01) | 0.16***<br>(0.01) | 0.15***<br>(0.01) | 0.08***<br>(0.01) | -0.08***<br>(0.01) | 0.13***<br>(0.01) | 0.07***<br>(0.01) | 0.06***<br>(0.01) | 0.06***<br>(0.01) |

\*\*\* $p < 0.001$ ; \*\* $p < 0.01$ ; \* $p < 0.05$
\*\*\* $p < 0.001$ ; \*\* $p < 0.01$ ; \* $p < 0.05$

**Table 8:** Logistic Regression Results for Age Categories. The dependent variable is the Forced Choice decision. These are the estimates used to construct the conjoint plots presented in Figure 2.

|  | Age |  |  |
| --- | --- | --- | --- |
|  | 18-39 | 40-59 | 60+ |
| (Intercept) | -1.19***<br>(0.10) | -1.41***<br>(0.10) | -1.72***<br>(0.13) |
| Vulnerability |  |  |  |
| Moderate (risk of death) | 0.23***<br>(0.04) | 0.22***<br>(0.04) | 0.19***<br>(0.04) |
| High (risk of death) | 0.74***<br>(0.08) | 0.82***<br>(0.07) | 0.86***<br>(0.06) |
| Transmission |  |  |  |
| Moderate risk (of transmission) | 0.20***<br>(0.02) | 0.18***<br>(0.03) | 0.21***<br>(0.04) |
| High risk (of transmission) | 0.65***<br>(0.05) | 0.73***<br>(0.05) | 0.81***<br>(0.05) |
| Income |  |  |  |
| Average income level | -0.12***<br>(0.02) | -0.05*<br>(0.02) | -0.05*<br>(0.02) |
| Highest 20% income level | -0.24***<br>(0.04) | -0.19***<br>(0.05) | -0.18***<br>(0.03) |
| Occupation |  |  |  |
| Non-Key worker: Can work at home | -0.03<br>(0.05) | -0.04<br>(0.05) | 0.00<br>(0.05) |
| Non-Key worker: Cannot work at home | 0.34***<br>(0.05) | 0.39***<br>(0.05) | 0.38***<br>(0.04) |
| Key worker: Education and childcare | 0.80***<br>(0.07) | 0.92***<br>(0.07) | 1.05***<br>(0.07) |
| Key worker: Factory worker | 0.55***<br>(0.04) | 0.60***<br>(0.05) | 0.75***<br>(0.06) |
| Key worker: Water and electricity service | 0.58***<br>(0.05) | 0.66***<br>(0.06) | 0.87***<br>(0.07) |
| Key worker: Police and fire-fighting | 0.73***<br>(0.07) | 0.92***<br>(0.07) | 1.23***<br>(0.09) |
| Key worker: Health and social care | 1.01***<br>(0.08) | 1.16***<br>(0.08) | 1.44***<br>(0.08) |
| Age Category |  |  |  |
| 40 years old | 0.19***<br>(0.03) | 0.23***<br>(0.02) | 0.28***<br>(0.05) |
| 65 years old | 0.31***<br>(0.05) | 0.45***<br>(0.06) | 0.63***<br>(0.06) |
| 79 years old | 0.31***<br>(0.07) | 0.39***<br>(0.08) | 0.59***<br>(0.09) |
| Country Fixed Effects? | Yes | Yes | Yes |

\*\*\* $p < 0.001$ ; \*\* $p < 0.01$ ; \* $p < 0.05$

**Table 9:** Linear (OLS) Regression Results for Age Categories. The dependent variable is the Forced Choice decision. These are the estimates used to construct the conjoint plots presented in Figure 10.

|  | Age |  |  |
| --- | --- | --- | --- |
|  | 18-39 | 40-59 | 60+ |
| (Intercept) | 0.22***<br>(0.02) | 0.18***<br>(0.02) | 0.12***<br>(0.02) |
| Vulnerability |  |  |  |
| Moderate (risk of death) | 0.05***<br>(0.01) | 0.05***<br>(0.01) | 0.04***<br>(0.01) |
| High (risk of death) | 0.17***<br>(0.02) | 0.19***<br>(0.01) | 0.19***<br>(0.01) |
| Transmission |  |  |  |
| Moderate risk (of transmission) | 0.05***<br>(0.01) | 0.04***<br>(0.01) | 0.05***<br>(0.01) |
| High risk (of transmission) | 0.15***<br>(0.01) | 0.17***<br>(0.01) | 0.18***<br>(0.01) |
| Income |  |  |  |
| Average income level | -0.03***<br>(0.01) | -0.01*<br>(0.01) | -0.01*<br>(0.00) |
| Highest 20% income level | -0.06***<br>(0.01) | -0.04***<br>(0.01) | -0.04***<br>(0.01) |
| Occupation |  |  |  |
| Non-Key worker: Can work at home | -0.01<br>(0.01) | -0.01<br>(0.01) | 0.00<br>(0.01) |
| Non-Key worker: Cannot work at home | 0.08***<br>(0.01) | 0.09***<br>(0.01) | 0.08***<br>(0.01) |
| Key worker: Education and childcare | 0.19***<br>(0.02) | 0.21***<br>(0.01) | 0.24***<br>(0.01) |
| Key worker: Factory worker | 0.13***<br>(0.01) | 0.14***<br>(0.01) | 0.17***<br>(0.01) |
| Key worker: Water and electricity service | 0.14***<br>(0.01) | 0.15***<br>(0.01) | 0.20***<br>(0.01) |
| Key worker: Police and fire-fighting | 0.17***<br>(0.01) | 0.21***<br>(0.01) | 0.28***<br>(0.02) |
| Key worker: Health and social care | 0.24***<br>(0.02) | 0.27***<br>(0.02) | 0.32***<br>(0.01) |
| Age Category |  |  |  |
| 40 years old | 0.04***<br>(0.01) | 0.05***<br>(0.01) | 0.06***<br>(0.01) |
| 65 years old | 0.07***<br>(0.01) | 0.10***<br>(0.01) | 0.14***<br>(0.01) |
| 79 years old | 0.07***<br>(0.02) | 0.09***<br>(0.02) | 0.13***<br>(0.02) |
| Country Fixed Effects? | Yes | Yes | Yes |

\*\*\* $p < 0.001$ ; \*\* $p < 0.01$ ; \* $p < 0.05$

**Table 10:** Logistic Regression Results for Income Categories. The dependent variable is the Forced Choice decision. These are the estimates used to construct the conjoint plots presented in Figure 2.

|  | Income |  |
| --- | --- | --- |
|  | Low | High |
| (Intercept) | -1.31***<br>(0.14) | -1.46***<br>(0.09) |
| Vulnerability |  |  |
| Moderate (risk of death) | 0.21***<br>(0.04) | 0.25***<br>(0.04) |
| High (risk of death) | 0.75***<br>(0.08) | 0.88***<br>(0.05) |
| Transmission |  |  |
| Moderate risk (of transmission) | 0.19***<br>(0.02) | 0.21***<br>(0.02) |
| High risk (of transmission) | 0.68***<br>(0.06) | 0.74***<br>(0.04) |
| Income |  |  |
| Average income level | -0.09***<br>(0.02) | -0.05*<br>(0.02) |
| Highest 20% income level | -0.23***<br>(0.04) | -0.17***<br>(0.04) |
| Occupation |  |  |
| Non-Key worker: Can work at home | -0.06<br>(0.06) | 0.05<br>(0.03) |
| Non-Key worker: Cannot work at home | 0.33***<br>(0.06) | 0.40***<br>(0.03) |
| Key worker: Education and childcare | 0.83***<br>(0.09) | 0.96***<br>(0.05) |
| Key worker: Factory worker | 0.58***<br>(0.06) | 0.65***<br>(0.03) |
| Key worker: Water and electricity service | 0.63***<br>(0.07) | 0.73***<br>(0.05) |
| Key worker: Police and fire-fighting | 0.82***<br>(0.09) | 0.99***<br>(0.04) |
| Key worker: Health and social care | 1.10***<br>(0.10) | 1.18***<br>(0.09) |
| Age Category |  |  |
| 40 years old | 0.23***<br>(0.03) | 0.21***<br>(0.03) |
| 65 years old | 0.45***<br>(0.05) | 0.36***<br>(0.09) |
| 79 years old | 0.44***<br>(0.06) | 0.29*<br>(0.12) |
| Country Fixed Effects? | Yes | Yes |

\*\*\* $p < 0.001$ ; \*\* $p < 0.01$ ; \* $p < 0.05$

**Table 11:** Linear (OLS) Regression Results for Income Categories. The dependent variable is the Forced Choice decision. These are the estimates used to construct the conjoint plots presented in Figure 10.

|  | Income |  |
| --- | --- | --- |
|  | Low | High |
| (Intercept) | 0.20***<br>(0.03) | 0.16***<br>(0.02) |
| Vulnerability |  |  |
| Moderate (risk of death) | 0.05***<br>(0.01) | 0.06***<br>(0.01) |
| High (risk of death) | 0.17***<br>(0.02) | 0.20***<br>(0.01) |
| Transmission |  |  |
| Moderate risk (of transmission) | 0.04***<br>(0.00) | 0.05***<br>(0.00) |
| High risk (of transmission) | 0.16***<br>(0.01) | 0.17***<br>(0.01) |
| Income |  |  |
| Average income level | -0.02***<br>(0.00) | -0.01*<br>(0.01) |
| Highest 20% income level | -0.05***<br>(0.01) | -0.04***<br>(0.01) |
| Occupation |  |  |
| Non-Key worker: Can work at home | -0.01<br>(0.01) | 0.01<br>(0.01) |
| Non-Key worker: Cannot work at home | 0.08***<br>(0.01) | 0.09***<br>(0.01) |
| Key worker: Education and childcare | 0.19***<br>(0.02) | 0.22***<br>(0.01) |
| Key worker: Factory worker | 0.13***<br>(0.01) | 0.15***<br>(0.01) |
| Key worker: Water and electricity service | 0.15***<br>(0.02) | 0.17***<br>(0.01) |
| Key worker: Police and fire-fighting | 0.19***<br>(0.02) | 0.23***<br>(0.01) |
| Key worker: Health and social care | 0.26***<br>(0.02) | 0.27***<br>(0.02) |
| Age Category |  |  |
| 40 years old | 0.05***<br>(0.01) | 0.05***<br>(0.01) |
| 65 years old | 0.10***<br>(0.01) | 0.08***<br>(0.02) |
| 79 years old | 0.10***<br>(0.01) | 0.07*<br>(0.03) |
| Country Fixed Effects? | Yes | Yes |

\*\*\* $p < 0.001$ ; \*\* $p < 0.01$ ; \* $p < 0.05$

**Table 12:** Logistic Regression Results for Ideology Categories. The dependent variable is the Forced Choice decision. These are the estimates used to construct the conjoint plots presented in Figure 2.

|  | Ideology |  |
| --- | --- | --- |
|  | Left/Centre | Right |
| (Intercept) | -1.57***<br>(0.13) | -1.19***<br>(0.13) |
| Vulnerability |  |  |
| Moderate (risk of death) | 0.24***<br>(0.04) | 0.15***<br>(0.03) |
| High (risk of death) | 0.92***<br>(0.08) | 0.63***<br>(0.08) |
| Transmission |  |  |
| Moderate risk (of transmission) | 0.25***<br>(0.02) | 0.12***<br>(0.03) |
| High risk (of transmission) | 0.86***<br>(0.05) | 0.55***<br>(0.05) |
| Income |  |  |
| Average income level | -0.14***<br>(0.02) | -0.03<br>(0.03) |
| Highest 20% income level | -0.32***<br>(0.04) | -0.12**<br>(0.04) |
| Occupation |  |  |
| Non-Key worker: Can work at home | -0.07<br>(0.06) | 0.03<br>(0.02) |
| Non-Key worker: Cannot work at home | 0.41***<br>(0.06) | 0.31***<br>(0.05) |
| Key worker: Education and childcare | 1.04***<br>(0.06) | 0.75***<br>(0.09) |
| Key worker: Factory worker | 0.70***<br>(0.05) | 0.51***<br>(0.07) |
| Key worker: Water and electricity service | 0.78***<br>(0.06) | 0.56***<br>(0.09) |
| Key worker: Police and fire-fighting | 0.98***<br>(0.09) | 0.79***<br>(0.10) |
| Key worker: Health and social care | 1.38***<br>(0.07) | 0.97***<br>(0.10) |
| Age Category |  |  |
| 40 years old | 0.26***<br>(0.02) | 0.21***<br>(0.02) |
| 65 years old | 0.50***<br>(0.04) | 0.43***<br>(0.04) |
| 79 years old | 0.48***<br>(0.05) | 0.41***<br>(0.05) |
| Country Fixed Effects? | Yes | Yes |

\*\*\* $p < 0.001$ ; \*\* $p < 0.01$ ; \* $p < 0.05$

**Table 13:**
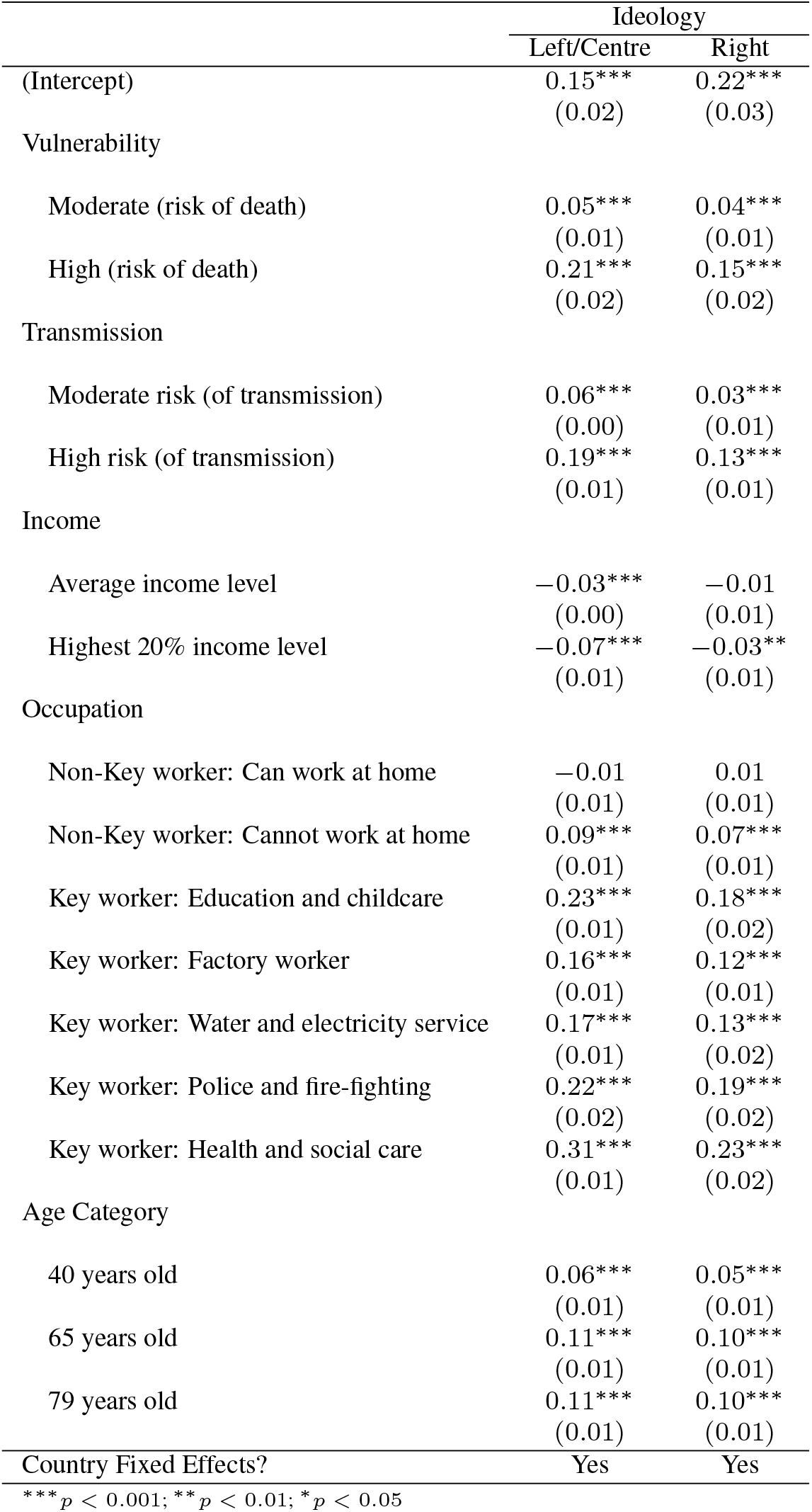
Linear (OLS) Regression Results for Ideology Categories. The dependent variable is the Forced Choice decision. These are the estimates used to construct the conjoint plots presented in Figure 10.

|  | Ideology |  |
| --- | --- | --- |
|  | Left/Centre | Right |
| (Intercept) | 0.15***<br>(0.02) | 0.22***<br>(0.03) |
| Vulnerability |  |  |
| Moderate (risk of death) | 0.05***<br>(0.01) | 0.04***<br>(0.01) |
| High (risk of death) | 0.21***<br>(0.02) | 0.15***<br>(0.02) |
| Transmission |  |  |
| Moderate risk (of transmission) | 0.06***<br>(0.00) | 0.03***<br>(0.01) |
| High risk (of transmission) | 0.19***<br>(0.01) | 0.13***<br>(0.01) |
| Income |  |  |
| Average income level | -0.03***<br>(0.00) | -0.01<br>(0.01) |
| Highest 20% income level | -0.07***<br>(0.01) | -0.03**<br>(0.01) |
| Occupation |  |  |
| Non-Key worker: Can work at home | -0.01<br>(0.01) | 0.01<br>(0.01) |
| Non-Key worker: Cannot work at home | 0.09***<br>(0.01) | 0.07***<br>(0.01) |
| Key worker: Education and childcare | 0.23***<br>(0.01) | 0.18***<br>(0.02) |
| Key worker: Factory worker | 0.16***<br>(0.01) | 0.12***<br>(0.01) |
| Key worker: Water and electricity service | 0.17***<br>(0.01) | 0.13***<br>(0.02) |
| Key worker: Police and fire-fighting | 0.22***<br>(0.02) | 0.19***<br>(0.02) |
| Key worker: Health and social care | 0.31***<br>(0.01) | 0.23***<br>(0.02) |
| Age Category |  |  |
| 40 years old | 0.06***<br>(0.01) | 0.05***<br>(0.01) |
| 65 years old | 0.11***<br>(0.01) | 0.10***<br>(0.01) |
| 79 years old | 0.11***<br>(0.01) | 0.10***<br>(0.01) |
| Country Fixed Effects? | Yes | Yes |

\*\*\* $p < 0.001$ ; \*\* $p < 0.01$ ; \* $p < 0.05$

**Table 14:** Logistic Regression Results for COVID-19 Vaccine Hesistancy Categories. The dependent variable is the Forced Choice decision. These are the estimates used to construct the conjoint plots presented in Figure 7.

|  | COVID-19 Vaccine Hesitancy |  |  |
| --- | --- | --- | --- |
|  | High | Moderate | Low |
| (Intercept) | -1.34***<br>(0.11) | -1.31***<br>(0.10) | -1.58***<br>(0.13) |
| Vulnerability |  |  |  |
| Moderate (risk of death) | 0.20***<br>(0.04) | 0.24***<br>(0.03) | 0.30***<br>(0.05) |
| High (risk of death) | 0.77***<br>(0.07) | 0.74***<br>(0.06) | 0.97***<br>(0.09) |
| Transmission |  |  |  |
| Moderate risk (of transmission) | 0.19***<br>(0.02) | 0.20***<br>(0.02) | 0.24***<br>(0.04) |
| High risk (of transmission) | 0.72***<br>(0.05) | 0.64***<br>(0.04) | 0.78***<br>(0.05) |
| Income |  |  |  |
| Average income level | -0.09***<br>(0.02) | -0.06<br>(0.03) | -0.09**<br>(0.03) |
| Highest 20% income level | -0.23***<br>(0.04) | -0.15**<br>(0.05) | -0.21***<br>(0.03) |
| Occupation |  |  |  |
| Non-Key worker: Can work at home | -0.05<br>(0.05) | 0.02<br>(0.04) | 0.05<br>(0.05) |
| Non-Key worker: Cannot work at home | 0.38***<br>(0.04) | 0.27***<br>(0.04) | 0.41***<br>(0.05) |
| Key worker: Education and childcare | 0.89***<br>(0.07) | 0.83***<br>(0.05) | 0.98***<br>(0.08) |
| Key worker: Factory worker | 0.60***<br>(0.05) | 0.59***<br>(0.03) | 0.66***<br>(0.05) |
| Key worker: Water and electricity service | 0.65***<br>(0.06) | 0.63***<br>(0.05) | 0.76***<br>(0.05) |
| Key worker: Police and fire-fighting | 0.86***<br>(0.08) | 0.93***<br>(0.06) | 1.00***<br>(0.08) |
| Key worker: Health and social care | 1.14***<br>(0.08) | 1.00***<br>(0.09) | 1.29***<br>(0.09) |
| Age Category |  |  |  |
| 40 years old | 0.23***<br>(0.03) | 0.21***<br>(0.04) | 0.20***<br>(0.05) |
| 65 years old | 0.41***<br>(0.06) | 0.38***<br>(0.08) | 0.47***<br>(0.07) |
| 79 years old | 0.39***<br>(0.07) | 0.34**<br>(0.11) | 0.46***<br>(0.11) |
| Country Fixed Effects? | Yes | Yes | Yes |

\*\*\* $p < 0.001$ ; \*\* $p < 0.01$ ; \* $p < 0.05$

**Table 15:** Linear (OLS) Regression Results for COVID-19 Vaccine Hesistancy Categories. The dependent variable is the Forced Choice decision. These are the estimates used to construct the conjoint plots presented in Figure 8.

|  | COVID-19 Vaccine Hesitancy |  |  |
| --- | --- | --- | --- |
|  | High | Moderate | Low |
| (Intercept) | 0.19***<br>(0.02) | 0.19***<br>(0.02) | 0.14***<br>(0.03) |
| Vulnerability |  |  |  |
| Moderate (risk of death) | 0.05***<br>(0.01) | 0.06***<br>(0.01) | 0.07***<br>(0.01) |
| High (risk of death) | 0.18***<br>(0.01) | 0.17***<br>(0.01) | 0.22***<br>(0.02) |
| Transmission |  |  |  |
| Moderate risk (of transmission) | 0.04***<br>(0.00) | 0.05***<br>(0.00) | 0.05***<br>(0.01) |
| High risk (of transmission) | 0.17***<br>(0.01) | 0.15***<br>(0.01) | 0.18***<br>(0.01) |
| Income |  |  |  |
| Average income level | -0.02***<br>(0.00) | -0.01<br>(0.01) | -0.02**<br>(0.01) |
| Highest 20% income level | -0.05***<br>(0.01) | -0.03**<br>(0.01) | -0.05***<br>(0.01) |
| Occupation |  |  |  |
| Non-Key worker: Can work at home | -0.01<br>(0.01) | 0.01<br>(0.01) | 0.01<br>(0.01) |
| Non-Key worker: Cannot work at home | 0.09***<br>(0.01) | 0.06***<br>(0.01) | 0.09***<br>(0.01) |
| Key worker: Education and childcare | 0.21***<br>(0.01) | 0.19***<br>(0.01) | 0.22***<br>(0.02) |
| Key worker: Factory worker | 0.14***<br>(0.01) | 0.14***<br>(0.01) | 0.15***<br>(0.01) |
| Key worker: Water and electricity service | 0.15***<br>(0.01) | 0.15***<br>(0.01) | 0.17***<br>(0.01) |
| Key worker: Police and fire-fighting | 0.20***<br>(0.02) | 0.22***<br>(0.01) | 0.23***<br>(0.02) |
| Key worker: Health and social care | 0.26***<br>(0.02) | 0.24***<br>(0.02) | 0.29***<br>(0.02) |
| Age Category |  |  |  |
| 40 years old | 0.05***<br>(0.01) | 0.05***<br>(0.01) | 0.05***<br>(0.01) |
| 65 years old | 0.09***<br>(0.01) | 0.09***<br>(0.02) | 0.10***<br>(0.01) |
| 79 years old | 0.09***<br>(0.02) | 0.08**<br>(0.03) | 0.10***<br>(0.02) |
| Country Fixed Effects? | Yes | Yes | Yes |

\*\*\* $p < 0.001$ ; \*\* $p < 0.01$ ; \* $p < 0.05$

**Table 16:**
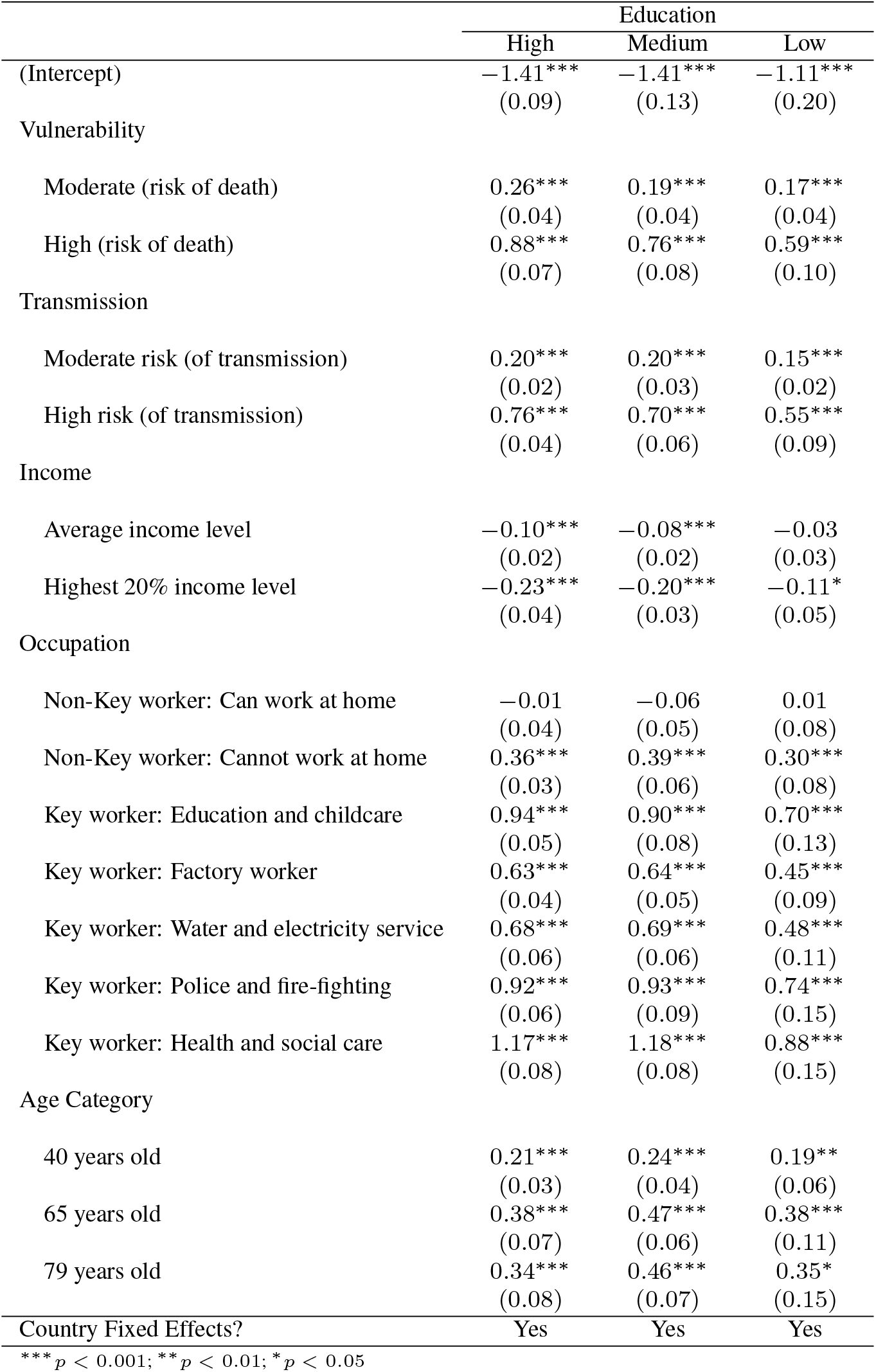
Logistic Regression Results for Education Categories. The dependent variable is the Forced Choice decision. These are the estimates used to construct the conjoint plots presented in Figure 7.

|  | Education |  |  |
| --- | --- | --- | --- |
|  | High | Medium | Low |
| (Intercept) | -1.41***<br>(0.09) | -1.41***<br>(0.13) | -1.11***<br>(0.20) |
| Vulnerability |  |  |  |
| Moderate (risk of death) | 0.26***<br>(0.04) | 0.19***<br>(0.04) | 0.17***<br>(0.04) |
| High (risk of death) | 0.88***<br>(0.07) | 0.76***<br>(0.08) | 0.59***<br>(0.10) |
| Transmission |  |  |  |
| Moderate risk (of transmission) | 0.20***<br>(0.02) | 0.20***<br>(0.03) | 0.15***<br>(0.02) |
| High risk (of transmission) | 0.76***<br>(0.04) | 0.70***<br>(0.06) | 0.55***<br>(0.09) |
| Income |  |  |  |
| Average income level | -0.10***<br>(0.02) | -0.08***<br>(0.02) | -0.03<br>(0.03) |
| Highest 20% income level | -0.23***<br>(0.04) | -0.20***<br>(0.03) | -0.11*<br>(0.05) |
| Occupation |  |  |  |
| Non-Key worker: Can work at home | -0.01<br>(0.04) | -0.06<br>(0.05) | 0.01<br>(0.08) |
| Non-Key worker: Cannot work at home | 0.36***<br>(0.03) | 0.39***<br>(0.06) | 0.30***<br>(0.08) |
| Key worker: Education and childcare | 0.94***<br>(0.05) | 0.90***<br>(0.08) | 0.70***<br>(0.13) |
| Key worker: Factory worker | 0.63***<br>(0.04) | 0.64***<br>(0.05) | 0.45***<br>(0.09) |
| Key worker: Water and electricity service | 0.68***<br>(0.06) | 0.69***<br>(0.06) | 0.48***<br>(0.11) |
| Key worker: Police and fire-fighting | 0.92***<br>(0.06) | 0.93***<br>(0.09) | 0.74***<br>(0.15) |
| Key worker: Health and social care | 1.17***<br>(0.08) | 1.18***<br>(0.08) | 0.88***<br>(0.15) |
| Age Category |  |  |  |
| 40 years old | 0.21***<br>(0.03) | 0.24***<br>(0.04) | 0.19**<br>(0.06) |
| 65 years old | 0.38***<br>(0.07) | 0.47***<br>(0.06) | 0.38***<br>(0.11) |
| 79 years old | 0.34***<br>(0.08) | 0.46***<br>(0.07) | 0.35*<br>(0.15) |
| Country Fixed Effects? | Yes | Yes | Yes |

\*\*\* $p < 0.001$ ; \*\* $p < 0.01$ ; \* $p < 0.05$

**Table 17:** Linear (OLS) Regression Results for Education Categories. The dependent variable is the Forced Choice decision. These are the estimates used to construct the conjoint plots presented in Figure 8.

|  | Education |  |  |
| --- | --- | --- | --- |
|  | High | Medium | Low |
| (Intercept) | 0.18***<br>(0.02) | 0.18***<br>(0.03) | 0.23***<br>(0.04) |
| Vulnerability |  |  |  |
| Moderate (risk of death) | 0.06***<br>(0.01) | 0.04***<br>(0.01) | 0.04***<br>(0.01) |
| High (risk of death) | 0.20***<br>(0.02) | 0.17***<br>(0.02) | 0.14***<br>(0.02) |
| Transmission |  |  |  |
| Moderate risk (of transmission) | 0.05***<br>(0.01) | 0.05***<br>(0.01) | 0.04***<br>(0.01) |
| High risk (of transmission) | 0.17***<br>(0.01) | 0.16***<br>(0.01) | 0.13***<br>(0.02) |
| Income |  |  |  |
| Average income level | -0.02***<br>(0.01) | -0.02***<br>(0.00) | -0.01<br>(0.01) |
| Highest 20% income level | -0.05***<br>(0.01) | -0.05***<br>(0.01) | -0.03*<br>(0.01) |
| Occupation |  |  |  |
| Non-Key worker: Can work at home | -0.00<br>(0.01) | -0.01<br>(0.01) | 0.00<br>(0.02) |
| Non-Key worker: Cannot work at home | 0.08***<br>(0.01) | 0.09***<br>(0.01) | 0.07***<br>(0.02) |
| Key worker: Education and childcare | 0.22***<br>(0.01) | 0.21***<br>(0.02) | 0.17***<br>(0.03) |
| Key worker: Factory worker | 0.14***<br>(0.01) | 0.15***<br>(0.01) | 0.11***<br>(0.02) |
| Key worker: Water and electricity service | 0.16***<br>(0.01) | 0.16***<br>(0.01) | 0.12***<br>(0.02) |
| Key worker: Police and fire-fighting | 0.21***<br>(0.01) | 0.22***<br>(0.02) | 0.18***<br>(0.03) |
| Key worker: Health and social care | 0.27***<br>(0.02) | 0.27***<br>(0.01) | 0.21***<br>(0.03) |
| Age Category |  |  |  |
| 40 years old | 0.05***<br>(0.01) | 0.06***<br>(0.01) | 0.04**<br>(0.01) |
| 65 years old | 0.09***<br>(0.01) | 0.11***<br>(0.01) | 0.09***<br>(0.03) |
| 79 years old | 0.08***<br>(0.02) | 0.10***<br>(0.02) | 0.08*<br>(0.03) |
| Country Fixed Effects? | Yes | Yes | Yes |

\*\*\* $p < 0.001$ ; \*\* $p < 0.01$ ; \* $p < 0.05$

**Table 18:** Logistic Regression Results for Gender Categories. The dependent variable is the Forced Choice decision. These are the estimates used to construct the conjoint plots presented in Figure 7.

|  | Gender |  |  |
| --- | --- | --- | --- |
|  | Female | Male | Other |
| (Intercept) | -1.49***<br>(0.13) | -1.24***<br>(0.10) | -1.16***<br>(0.29) |
| Vulnerability |  |  |  |
| Moderate (risk of death) | 0.25***<br>(0.04) | 0.19***<br>(0.03) | 0.35<br>(0.33) |
| High (risk of death) | 0.86***<br>(0.07) | 0.72***<br>(0.07) | 1.01***<br>(0.22) |
| Transmission |  |  |  |
| Moderate risk (of transmission) | 0.22***<br>(0.03) | 0.17***<br>(0.02) | 0.03<br>(0.16) |
| High risk (of transmission) | 0.78***<br>(0.06) | 0.63***<br>(0.05) | 0.67**<br>(0.23) |
| Income |  |  |  |
| Average income level | -0.10***<br>(0.02) | -0.06***<br>(0.02) | -0.21<br>(0.22) |
| Highest 20% income level | -0.24***<br>(0.05) | -0.18***<br>(0.03) | -0.57***<br>(0.16) |
| Occupation |  |  |  |
| Non-Key worker: Can work at home | -0.02<br>(0.05) | -0.04<br>(0.04) | 0.46<br>(0.34) |
| Non-Key worker: Cannot work at home | 0.44***<br>(0.06) | 0.29***<br>(0.04) | 0.91*<br>(0.46) |
| Key worker: Education and childcare | 0.99***<br>(0.07) | 0.79***<br>(0.07) | 1.91***<br>(0.34) |
| Key worker: Factory worker | 0.69***<br>(0.06) | 0.52***<br>(0.04) | 1.45***<br>(0.33) |
| Key worker: Water and electricity service | 0.72***<br>(0.06) | 0.60***<br>(0.06) | 1.49***<br>(0.32) |
| Key worker: Police and fire-fighting | 0.99***<br>(0.08) | 0.79***<br>(0.08) | 1.13***<br>(0.33) |
| Key worker: Health and social care | 1.28***<br>(0.09) | 1.00***<br>(0.08) | 1.88***<br>(0.40) |
| Age Category |  |  |  |
| 40 years old | 0.22***<br>(0.02) | 0.22***<br>(0.04) | 0.14<br>(0.20) |
| 65 years old | 0.42***<br>(0.06) | 0.41***<br>(0.07) | 0.47<br>(0.28) |
| 79 years old | 0.41***<br>(0.07) | 0.37***<br>(0.09) | 0.47<br>(0.25) |
| Country Fixed Effects? | Yes | Yes | Yes |

\*\*\* $p < 0.001$ ; \*\* $p < 0.01$ ; \* $p < 0.05$

**Table 19:** Linear (OLS) Regression Results for Gender Categories. The dependent variable is the Forced Choice decision. These are the estimates used to construct the conjoint plots presented in Figure 8.

|  | Gender |  |  |
| --- | --- | --- | --- |
|  | Female | Male | Other |
| (Intercept) | 0.16***<br>(0.03) | 0.21***<br>(0.02) | 0.26***<br>(0.06) |
| Vulnerability |  |  |  |
| Moderate (risk of death) | 0.06***<br>(0.01) | 0.05***<br>(0.01) | 0.07<br>(0.07) |
| High (risk of death) | 0.20***<br>(0.01) | 0.17***<br>(0.01) | 0.22***<br>(0.04) |
| Transmission |  |  |  |
| Moderate risk (of transmission) | 0.05***<br>(0.01) | 0.04***<br>(0.00) | 0.01<br>(0.03) |
| High risk (of transmission) | 0.18***<br>(0.01) | 0.15***<br>(0.01) | 0.14**<br>(0.05) |
| Income |  |  |  |
| Average income level | -0.02***<br>(0.01) | -0.02***<br>(0.00) | -0.04<br>(0.05) |
| Highest 20% income level | -0.05***<br>(0.01) | -0.04***<br>(0.01) | -0.12***<br>(0.03) |
| Occupation |  |  |  |
| Non-Key worker: Can work at home | -0.00<br>(0.01) | -0.01<br>(0.01) | 0.09<br>(0.07) |
| Non-Key worker: Cannot work at home | 0.10***<br>(0.01) | 0.07***<br>(0.01) | 0.19*<br>(0.09) |
| Key worker: Education and childcare | 0.23***<br>(0.01) | 0.19***<br>(0.02) | 0.41***<br>(0.07) |
| Key worker: Factory worker | 0.16***<br>(0.01) | 0.12***<br>(0.01) | 0.31***<br>(0.07) |
| Key worker: Water and electricity service | 0.17***<br>(0.01) | 0.14***<br>(0.01) | 0.32***<br>(0.06) |
| Key worker: Police and fire-fighting | 0.23***<br>(0.02) | 0.19***<br>(0.02) | 0.24***<br>(0.07) |
| Key worker: Health and social care | 0.29***<br>(0.02) | 0.23***<br>(0.02) | 0.40***<br>(0.08) |
| Age Category |  |  |  |
| 40 years old | 0.05***<br>(0.00) | 0.05***<br>(0.01) | 0.03<br>(0.04) |
| 65 years old | 0.09***<br>(0.01) | 0.09***<br>(0.02) | 0.10<br>(0.06) |
| 79 years old | 0.09***<br>(0.02) | 0.09***<br>(0.02) | 0.10<br>(0.05) |
| Country Fixed Effects? | Yes | Yes | Yes |

\*\*\* $p < 0.001$ ; \*\* $p < 0.01$ ; \* $p < 0.05$

## Footnotes

1 In the Supplementary Materials, see Tables 2 for sample size details and Tables 4 and 5 for the distribution of covariates for each country.

2 Other recent policy-related illustrations of conjoint experiments include (*28*)

3 Figure 4 in Supplementary Materials provides an example of the attributes and values that characterized the two vaccine candidates presented to respondents. Checking the proportion of times individual conjoint levels were shown to subjects confirms that they were adequately randomised (see Supplement Table 3).

4 Identical models, using the same dichotomous outcome variable, were estimated using OLS regression with clustered standard errors. These results are reported in Supplementary Materials Table 7. Supplementary Materials Figure 6 displays the estimated average marginal component-specific effects (AMCEs) generated from the OLS regressions (*30*). The results are virtually identical to those presented in the main text.

5 The exact wording of these questions is reported in the Supplementary Materials.

6 Further information on Respondi is available here: https://www.respondi.com/EN/.

7 The recruitment ad is available in the Online Appendix.

8 Other recent policy-related illustrations of conjoint experiments include (*28*)

